# Auditory Network Discoherence in Chronic Tinnitus

**DOI:** 10.64898/2026.06.01.26354620

**Authors:** Amber M. Leaver

## Abstract

Chronic tinnitus is a common condition with few effective treatments and no cure. Though inconsistent results across MRI studies of tinnitus have slowed mechanistic insight, converging evidence across animal and human studies clearly implicate auditory-system dysfunction. This paper presents a systematic, retrospective assessment of auditory-network function in chronic tinnitus across multiple fMRI datasets. Auditory network nodes were newly defined in this effort, including novel nodes in cerebellum previously linked with somatotopic representations of articulators (lobules VI, VIIIa). Auditory-network connectivity in cerebellum and superior olivary complex was reduced in chronic tinnitus, perhaps explaining the recent success of trigeminal stimulation in improving tinnitus. Auditory-network strength was also reduced, corroborating some recent studies and perhaps reflecting increased spontaneous neuronal activity reported in animal models. Together, these results suggest auditory-network dysconnectivity as a tinnitus biomarker, and that efferent cochlear pathways related to head-centric interoception may play a mechanistic role.

## INTRODUCTION

A large number of MRI studies have measured how brain function differs in chronic tinnitus. These studies address a pressing need: to understand why tinnitus becomes chronic in some people but not others, even when exposed to the same conditions that appear to cause tinnitus like hearing/ear damage, ototoxicity, or stress. A mechanistic understanding of chronic tinnitus is sorely needed, given the ubiquity of the condition (10-15% of global population [1]) and lack of curative treatments. However, results across existing tinnitus fMRI studies are widely varied, likely due to challenges not unique to this field (e.g., small sample sizes, differing analysis techniques and field of view, multiple statistical comparisons [2]). This has lessened the translational impact of tinnitus fMRI studies and slowed mechanistic insight.

Despite these challenges, converging evidence across animal and human research clearly implicate auditory-system dysfunction in tinnitus. Chronic tinnitus is thought to be caused when input from the inner ear to the ascending auditory system is lost or altered due to hair cell damage and/or interference from the somatosensory system [3, 4]. In animal models of tinnitus, studies show increased spontaneous neuronal firing rates and sound-evoked hyperactivity within the ascending auditory pathway [4]. Functional MRI studies in people with chronic tinnitus have also reported increased sound-evoked activity within the ascending auditory pathway [5, 6], as well as decreased temporal coherence in resting-state fMRI data (i.e., functional connectivity) in and between ascending auditory regions [7–10]. Some have discussed potential links between this latter fMRI finding and increased spontaneous firing rates in animal models [10, 11]. Though there is debate regarding which part(s) of the ascending auditory pathway contribute to chronic tinnitus, there is consensus that some kind of aberrant auditory-system activity is key contributing factor [3, 4, 12–14]. Thus, discoherence in resting-state fMRI activity (i.e., decreased functional connectivity) could be a sensible biomarker for chronic tinnitus treatment studies.

Given this evidence of central auditory dysfunction in chronic tinnitus, there has been interest in targeting this dysfunction with noninvasive brain stimulation. Meta-analyses of transcranial magnetic stimulation (TMS) and transcranial direct current stimulation (tDCS) protocols targeting auditory cortex show a statistical advantage for active stimulation over sham stimulation [15, 16]. Yet, improvements are small and inconsistent across studies, making it unclear which stimulation protocols are most effective. In our recent pilot trial of focal auditory-cortex tDCS, volunteers rated their tinnitus as being ∼24% quieter after five tDCS sessions, a significant improvement over sham [17]. Identifying a measurable improvement in tinnitus after only five sessions is notable, given that this is far fewer than the 15-20 sessions used FDA-cleared clinical TMS protocols for depression, addiction, and OCD. Furthermore, auditory-network connectivity also increased after the first tDCS session in this protocol, perhaps providing indirect support for auditory-network discoherence as a potential tinnitus biomarker. Taken together, available evidence suggests that noninvasive brain stimulation protocols could be optimized to increase auditory-network coherence, perhaps making tinnitus quieter and/or less intrusive.

This study tested the hypotheses that auditory-network connectivity measured with fMRI is reduced, or “discoherent,” in chronic tinnitus, and that tDCS protocols targeting auditory cortex can normalize this discoherence. A series of retrospective analyses attempted to identify consistent effects across resting-state fMRI analysis methods, including seed- vs. network-based definitions of functional connectivity, node vs. network connectivity strength, and statistical control for potential confounding variables like age, hearing function, and depression. Though originally designed for well-controlled longitudinal analysis, these retrospective multi-cohort analyses offer greater potential sensitivity to detect small effect sizes due to larger sample sizes [18, 19] and include hearing assessments not included other large-scale MRI datasets like the Human Connectome Project [20, 21]. Auditory cortex was also stimulated noninvasively either during or immediately before fMRI using conventional/sponge or 4×1 Ag/AgCl electrodes, respectively. Controls for these tDCS studies included both sham and active stimulation. Taken together, the goal of these analyses was to better understand auditory network connectivity and its modulation with tDCS in chronic tinnitus.

## MATERIALS AND METHODS

### Source Data

Data analyzed in this study were combined from previously published MRI studies of tES of chronic tinnitus and depression (NCT04031547 & NCT03544359 [22], NCT05120037 [17], NCT05090397 [23]). As detailed below, these studies used near-identical MRI sequences and surveys of demographic, medical history, and behavioral data. Data are locally held by the author, but may be made available upon reasonable request meeting institutional regulatory standards.

### Participants

Participants were recruited from communities surrounding Northwestern University and University of California Los Angeles, and gave informed consent to participate in their original studies in accordance with the Declaration of Helsinki and with approval from the Institutional Review Board of each respective institution. These combined datasets included 117 participants (3 omitted after visual inspection of MRI data); 43 of these had chronic tinnitus, 35 had current or past clinical diagnosis of major depression, and 8 reported both chronic tinnitus and depression diagnosis. Demographic and other details are given in **Tables 1 and S1**. Participants with chronic tinnitus reported experiencing tinnitus for one year or longer, with no hyperacusis or Meniere’s disease. Exclusion criteria for all participants included MRI or tES contraindications (e.g., claustrophobia, certain implants), significant history of neuropsychiatric or neurologic condition, and significant past head injury. Detailed inclusion/exclusion criteria are listed in **Supplemental Methods**.

### Audiometry

Hearing function was measured using pure-tone thresholds in clinical and/or research settings. Clinical audiometry was performed at UCLA and Northwestern Medicine audiology clinics using calibrated equipment and in sound-proof booths. Research audiometry was performed using the Hearing Thresholds Test 7+ v1.0 from the NIH Toolbox 1.23.4940 using an iPad tablet (8^th^ generation) and Sennheiser Professional HD 280 PRO over-ear headphones. For research audiometry using NIH Toolbox, volume was kept at the same level for all participants (50% max), which yields pure-tone thresholds .2-8kHz similar to clinical pure-tone thresholds [24–26]. For both clinical and research audiometry, pure-tone threshold was averaged across both ears and all frequencies up to 8kHz to arrive at a single measure of hearing function/loss. Thirty participants had clinical audiometry only, 25 had research audiometry only, 21 had both, and 41 had neither. For participants with both, clinical pure-tone threshold averages were used.

### Tinnitus and Depression Assessments

Tinnitus impact was primarily assessed using the Tinnitus Handicap Inventory (missing for 3 participants) [27]. Tinnitus Functional Index served as a secondary measure of tinnitus impact (missing for 8 participants) [28]. Depression symptoms were assessed using the Beck Depression Inventory (21 item) in all participants.

### MRI Acquisition and Preprocessing

Resting state BOLD fMRI data were acquired using 3T Prisma scanners using identical sequences, including 2mm isotropic and 0.8s spatial and temporal resolution, respectively (details in **Supplement**). Data were analyzed using the FSL Feat tool, with boundary-based image registration (BBR) to single-subject anatomical data and MNI template [29]. Band-passed temporal filter was applied with cutoffs approximating 0.01 and 0.1 Hz, and images were smoothed at 4mm^3^. Motion was estimated using FSL’s mcflirt (as part of feat) and Motion Outliers tools using default settings, and these parameters were used in statistical analyses as described further below. NeuroCombat was used to mitigate cross-site differences in fMRI data [30] using R (https://www.r-project.org). NeuroCombat uses an empirical Bayesian approach originally developed to mitigate batch effects in genomics [31], and has been successfully applied to structural and functional MRI metrics in a variety of contexts [30, 32–35]. Freesurfer’s recon-all was applied to T1- and T2-weighted anatomical scans using standard parameters, in order to identify the location of Heschl’s (i.e., transverse temporal) gyrus and sulcus in each participant’s native space.

### Measuring Functional Connectivity: HG/HS Seed

A seed-based approach was the primary method used to assess auditory-network connectivity (i.e., temporal coherence in BOLD fMRI data). Bilateral Heschl’s Gyrus and Sulcus (HG/HS) were identified in each participant’s native space using Freesurfer Destrieux (aparc.2009s) atlas [36], and served as the seed region for these analyses. Resting-state BOLD fMRI timecourses were averaged within the bilateral HG/HS seed for each participant, and general linear models measured whether resting-state timecourses (explained variable) correlated with the bilateral HG/HS seed (regressor of interest) controlling for global signal, mean white matter signal, and motion (linear motion in six directions and outliers; nuisance regressors) using the fsl_glm command in FSL. The rationale for this seed-based approach was to emphasize activity in primary auditory cortical regions, which are most connected with the ascending auditory pathway and responsive to simple sounds similar to tinnitus percepts.

### Measuring Functional Connectivity: Dual Regression

A complementary measure of auditory-network connectivity used the standard Yeo17 resting state network atlas (17 networks liberal mask [37]) and FSL’s Dual Regression [38], to serve as a validation and complement to the seed-based approach. In brief, resting-state BOLD fMRI timecourses were averaged within each of the Yeo17 networks for each participant in MNI space, and general linear models identified voxels with resting-state timecourses (explained variable) that correlated with the Yeo17 auditory network (regressor of interest) controlling for the remaining 16 networks (nuisance regressors). Thus, the dual regression approach considers neurobiological resting-state signal from other non-auditory networks as nuisance regressors, while the seed-based approach does not. Other differences include using a standardized template vs. single-subject seed, network- vs. region-based seed definitions, and calculations in MNI vs. native space. It is important to note that the Yeo17 auditory network combines both auditory and ventral somatomotor cortices, but is nevertheless a useful comparator given its widespread use.

### Defining the Auditory Network (Auditory Network)

Statistical analyses used a node-based (i.e., region of interest) approach, with the goal of improving signal to noise and sensitivity to detect effects of tinnitus compared to voxelwise analyses. To define auditory-network nodes, voxelwise HG/HS seed maps from all participants were normalized to MNI template and averaged to create a voxelwise group map of the auditory network. Auditory network nodes were defined across a range of thresholds to include nodes with lower connectivity (e.g., ventral striatum) and to avoid overly large clusters (details in **Supplement**). Resulting nodes and associated thresholds are displayed in **Figure 1 and Table S2**. Primary auditory regions were defined anatomically using Jülich atlas [39, 40], and inferior colliculus (IC) was defined manually on the MNI template (IC; MNI_x,y,z_ = −5, −36.5, −10.5 for left IC and MNI_x,y,z_ = 6, −36.5, −10.5 for right). Post hoc analyses confirmed that mean node strength was greater than zero in each node (all p_FDR_ < 0.05, one sample t test). Network Strength was calculated as the mean functional connectivity across all nodes [41]. Network strength and all nodes were used in all statistical analyses.

**Figure 1.**
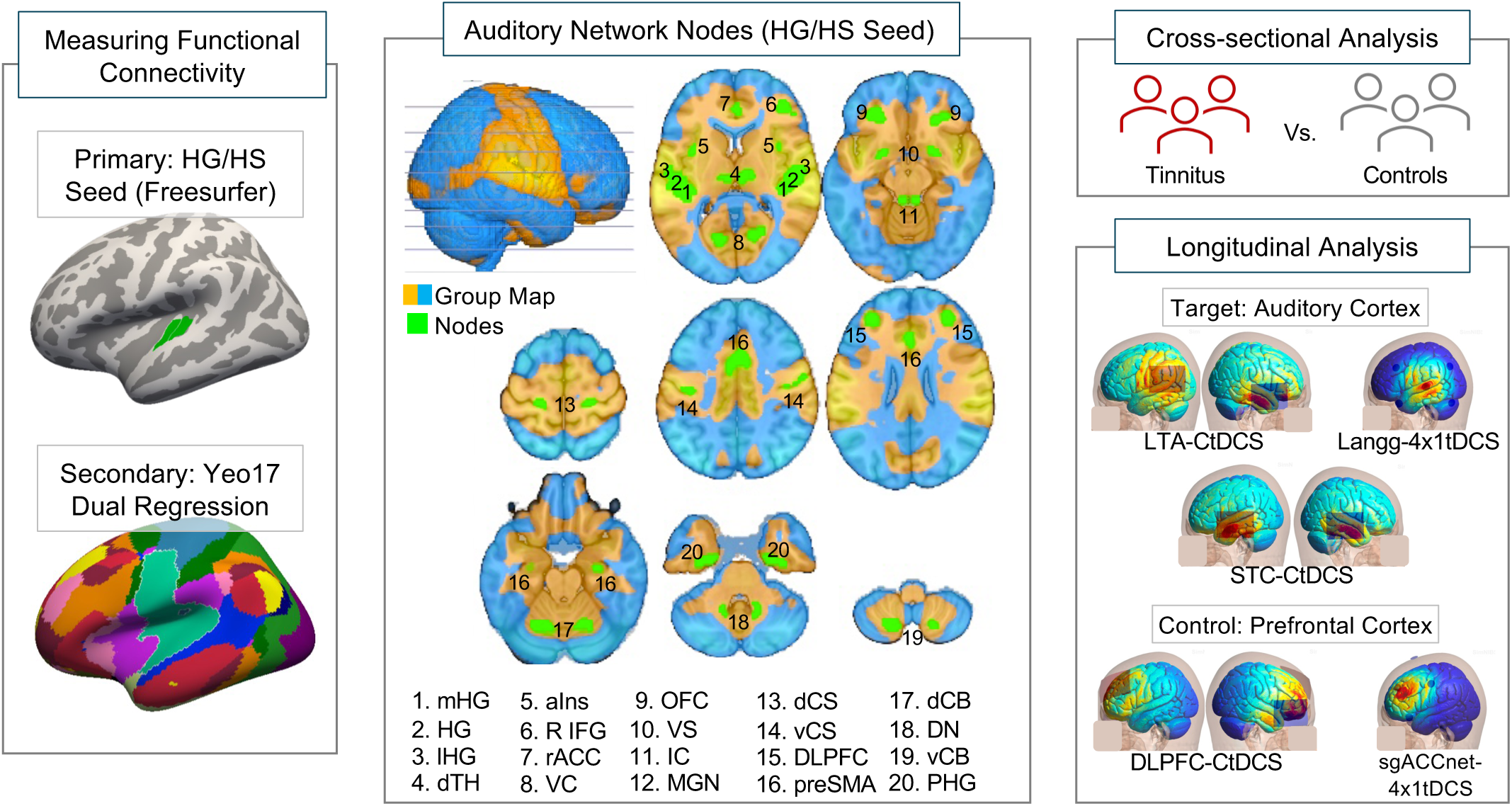
Methods summary for defining and measuring Auditory Network connectivity. First, a seed region comprising bilateral Heschl’s Gyrus & Sulcus (HG/HS) was defined in each participant’s native space in Freesurfer. Then, temporal coherence, i.e. functional connectivity, with the HG/HS seed was calculated voxelwise for each fMRI. Resulting native-space statistical maps were then warped to MNI space and averaged to create a group map of Auditory Network connectivity shown in center panel. Network Nodes (green) were positively correlated with the HG/HS timecourse (orange). HG/HS functional connectivity was averaged within these nodes for each scan, and subjected to cross-sectional analyses of tinnitus status and longitudinal analyses of tDCS effects.

### Statistical Analysis

#### Overview

Statistical analyses used R software [42]. Demographic, behavioral, and clinical variables were assessed for differences between groups (e.g., tinnitus vs. no tinnitus) using two-sided t tests, ANOVA, or chi-squared tests. Statistical analyses of auditory-network connectivity used linear mixed-effects models with participant as a random factor (i.e., across repeat fMRI scans) and appropriate nuisance factors (e.g., age, sex, MRI motion) as described further below.

False Discovery Rate (FDR) was estimated using the Benjamini-Hochberg method to correct for multiple comparisons across nodes. There are strong a priori hypotheses regarding auditory-system involvement in tinnitus; therefore, a restricted number of nodes/metrics were discussed at uncorrected p < 0.05 (Network Strength and auditory cortex). Standardized effect sizes were reported as available for each statistic, including semi-partial R squared (r2glmm [43]) and Cohen’s d (emmeans [44]).

### Statistical Analysis

#### Cross-sectional Study of Tinnitus Status

In the main cross-sectional analysis, linear mixed effects models assessed whether auditory-network connectivity (nodewise or Network Strength; explained variable) differed between people with and without tinnitus (factor of interest) controlling for participant (random factor), as well as age, sex, site, depression status, motion, visit, and tDCS condition (nuisance factors). The goal of this main cross-sectional model was to improve sensitivity to detect differences related to tinnitus by including as much data as possible, while controlling for as many relevant nuisance factors as possible. Because pure-tone thresholds were not available for all participants, this main model did not control for hearing function. A series of follow-up tests included mean bilateral pure-tone thresholds as a nuisance factor and (separately) excluded active, inactive, and post-tDCS fMRI sessions (**Table 2**).

Relationships between auditory-network connectivity (explained variable) and tinnitus impact (factor of interest) were also assessed using linear mixed effects models. These analyses used Tinnitus Handicap Inventory (THI) because these scores were present for the greatest number of tinnitus participants. This model controlled for participant (random factor) and age, sex, site, motion, visit, and tDCS condition (nuisance factors).

### Statistical Analysis

#### Longitudinal Study of tDCS

This next group of analyses compared active, sham, and rest tDCS conditions using the same source MRI data as described above. Study designs are described in source publications [17, 22, 26]. All studies included active, sham, and rest (i.e., no stimulation) conditions, with BOLD fMRI measured either online during tDCS or offline after tDCS. Statistical analyses used linear mixed effects models to assess changes in auditory-network connectivity (nodewise or Network Strength; explained variable) across tDCS conditions (factor of interest) controlling for participant (random factor), as well as age, sex, and other nuisance factors relevant to each study design described below.

Conventional/sponge (C) tDCS studies measured online effects of CtDCS during BOLD-fMRI scans. In 33 participants (16 tinnitus), anode was positioned over CP5 and cathode over FT8; this is a standard LTA (left temporoparietal area) montage used in previous tinnitus research. In 10 tinnitus participants, anode was positioned over T7 and cathode over T8 to target auditory cortex (superior temporal cortex, STC). In 37 (2 tinnitus) participants, anode was positioned over F3 and cathode over F8 in a standard montage used in previous depression research targeting left dorsolateral prefrontal cortex (DLPFC). Each CtDCS volunteer received 5 minutes of 2mA active or sham stimulation in randomized, counterbalanced order. Linear mixed effects models targeted a main effect of tDCS condition (active/sham/rest) controlling for participant (random factor) and age, sex, run, site, tinnitus (or depression) status, and motion (nuisance variables). Effects were considered significant if p_FDR_ < 0.05 for main effect (F statistic estimated using Type III sum of squares) and uncorrected p < 0.05 for active vs. sham conditions (emmeans).

In 4×1 tDCS studies, offline effects were measured, comparing BOLD-fMRI scans before and after stimulation. Small Ag/AgCl electrodes were placed in a 4×1 configuration (i.e., High Definition or HD, *Soterix*) with center anode and surround cathodes targeting either auditory cortex (20 tinnitus participants) or left dorsolateral prefrontal cortex (25 participants; 3 tinnitus). Each volunteer was randomized to receive 20 minutes of 2mA active or sham stimulation. MRI scans occurred immediately before and after the 20-minute stimulation session. An additional baseline MRI scan occurred approximately 1-2 weeks before the stimulation visit for the DLPFC study. Linear mixed effects models targeted an interaction between tDCS assignment (active/sham) and time (pre/post) controlling for participant (random factor) and age, sex, and motion (nuisance variables), separately for each montage. Effects meeting both the following criteria were considered significant (i.e., conjunction of both): p_FDR_ < 0.05 for interaction (t statistic, Type III sum of squares, LmerTest [45]) and uncorrected p < 0.05 for pre vs. post active condition (emmeans).

### Follow-up Analyses in Superior Olivary Complex (SOC)

After identifying effects of tinnitus status on auditory-network connectivity in cerebellar nodes, a follow-up analysis in superior olivary complex was explored. Group-averaged voxelwise maps of both HG/HS Seed and Dual Regression connectivity were inspected across thresholds (Supplementary Methods), and clusters in brainstem overlapping the position of the superior olivary complex were identified in the Dual Regression map. The position of these clusters were qualitatively confirmed using a structural atlas [46]. Statistical models were applied as described above for other nodes.

## RESULTS

### Identifying auditory network nodes

When assessing HG/HS seed-based functional connectivity, a series of brain regions were identified as nodes forming an auditory network (**Figure 1, Table S2**). Nodes identified in the ascending sensory pathway included auditory cortex and thalamus (i.e., medial geniculate nucleus, MGN). Auditory cortical regions were separated and inferior colliculus (IC) was identified using anatomical scans. Outside the ascending auditory system, cortical nodes were identified in prefrontal, dorsal anterior cingulate, and orbitofrontal cortex, anterior insula, presupplementary motor area, other sensory cortex (central sulcus, visual cortex), and parahippocampal cortex. Subcortical nodes were identified in dorsal thalamus, ventral striatum, cerebellum dentate nuclei, and cerebellum lobules VI and VIIIa. These nodes were also present when using the Yeo17 atlas and dual regression with some spatial differences.

### Reduced Auditory Network Strength in Tinnitus

In the main statistical model measuring HG/HS seed-based connectivity in the complete dataset, Auditory Network Strength was less in participants with chronic tinnitus (p_uncorr_ = 0.01, **Figure 2**). This effect was consistent across all follow-up tests, with tinnitus status explaining 3-7% of the variance in Auditory Network Strength across all models tested (i.e., r^2^_partial_ =0.03-0.07, **Table 2**). Whole-model fit explained 27% of Auditory Network Strength variance when including the complete dataset; additional statistics can be found in **Table S3** including for confound regressors. Of note, people with depression also showed reduced Auditory Network Strength in some statistical models (**Table S3**), though an interaction between depression and tinnitus status was not apparent (**Figure S1**).

**Figure 2.**
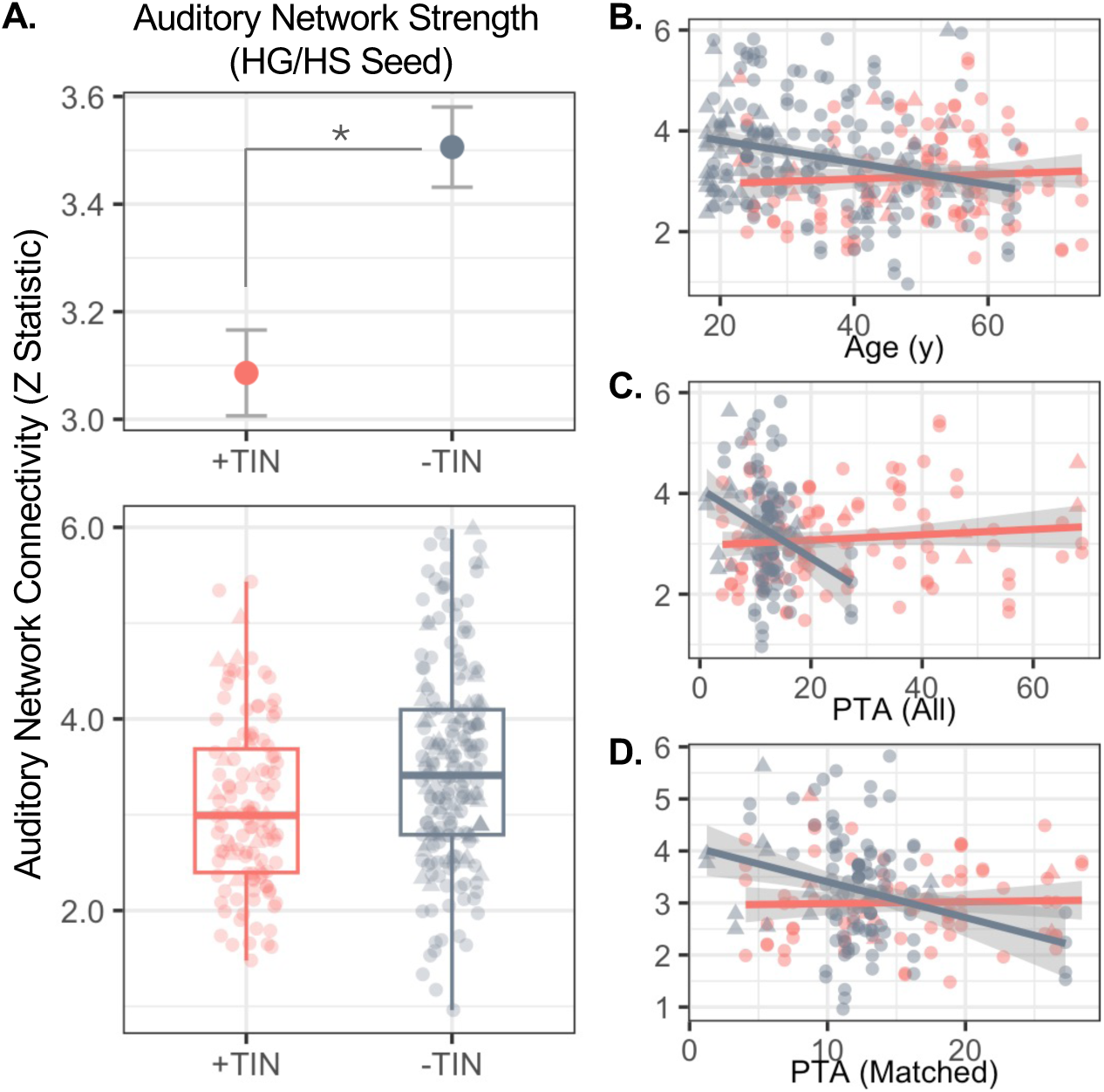
Auditory Network Strength was less in people with chronic tinnitus (+TIN, red) compared to controls (-TIN, gray). **A.** Upper panel displays mean and standard error, with corresponding boxplot below. **B-D.** Auditory Network Strength is plotted on the y axis against age, pure tone average (PTA) for frequencies <=8kHz for all participants, and PTA in a subset of participants with PTA < 30dB (Matched). Datapoints in A-D reflect data from single scans from NU and UCLA datasets in circles and triangles, respectively.

### Reduced Cerebellar Node Strength in Tinnitus

In the main statistical model measuring HG/HS Seed connectivity, functional connectivity in single nodes did not differ in tinnitus participants (p_FDR_ > 0.05). Though some auditory cortical nodes showed reduced connectivity strength in tinnitus participants, these effects did not reach chosen criteria for statistical significance (e.g., right HG r^2^_partial_ = 0.03, p_uncorr_=0.04; **Table S4**). However, when using the Dual Regression approach to statistically control for connectivity with other non-auditory resting-state networks in Yeo17 atlas, select cerebellar and dorsal central sulcus nodes showed less connectivity with the auditory network (p_FDR_ < 0.05; **Figure 3**, **Table S5**). Cerebellar nodes included left dorsal cerebellum (lobule VI) and dentate nucleus (e.g., left lobule VI r^2^_partial_ = 0.07 p_FDR_ = 0.021; **Table S5)**.

**Figure 3.**
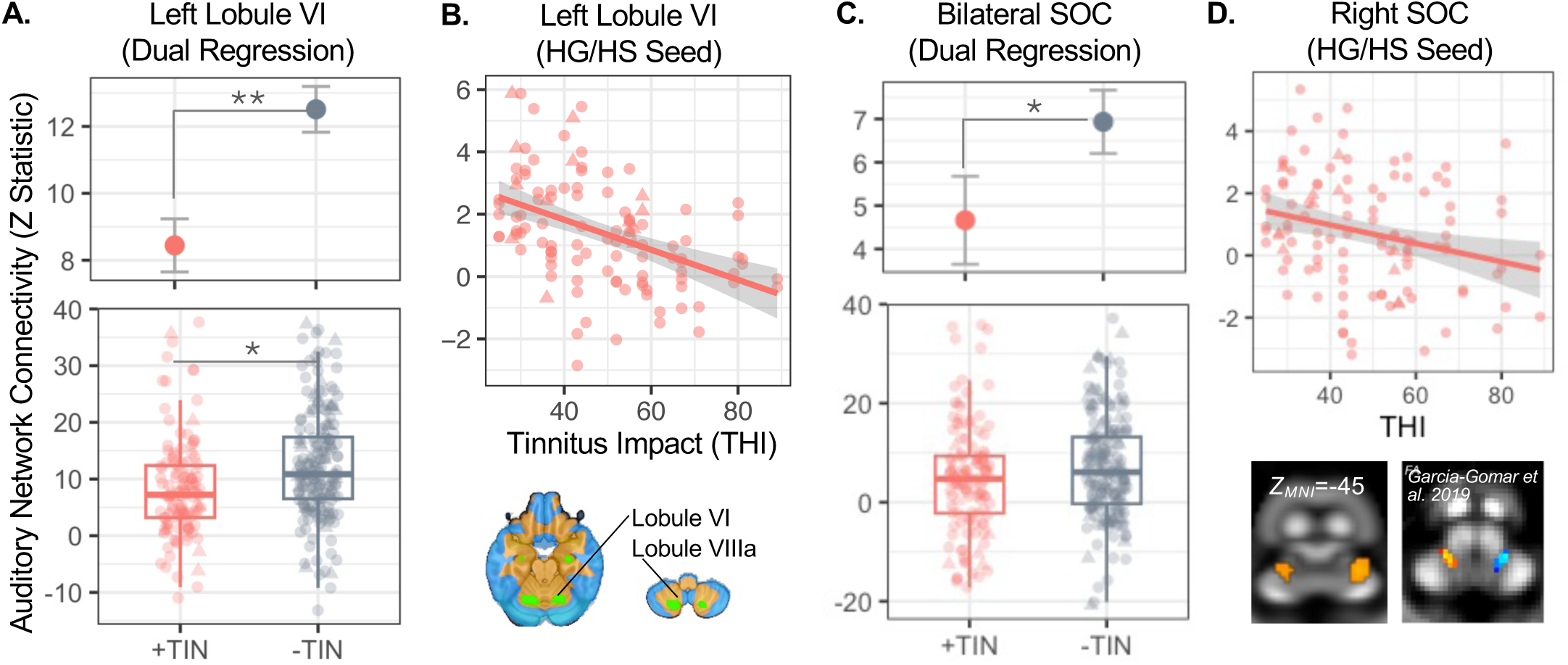
**A.** Auditory-network connectivity was less in cerebellum nodes for people with chronic tinnitus (+TIN, red) compared to controls (-TIN, gray) when using dual regression. Upper panel displays means and standard error, with corresponding boxplots in lower panel. **B.** Scatterplot displays the negative relationship between tinnitus impact (Tinnitus Handicap Inventory score) and auditory-network connectivity in left lobule VI measured using HG/HS Seed method. Inset below displays the location of relevant cerebellar nodes in green (see also Fig 1). **C.** Auditory-network connectivity was less in bilateral superior olivary complex in people with tinnitus; data are plotted as in A. **D.** Scatterplot displays negative correlation between right SOC connectivity and THI as in panel B. Inset below panel D shows position of SOC nodes overlaid on standard template white matter image (fractional anisotropy) in MNI space at left, which can be qualitatively compared image from in vivo atlas from García-Gomar et al. 2019 at right. Permission to reproduce this image is granted under the terms of Creative Commons Attribution License CC-BY. Datapoints in A-D reflect data from single scans from NU and UCLA datasets in circles and triangles, respectively.

Tinnitus impact (THI score) negatively correlated with left dorsal cerebellar (lobule VI) node strength when measuring HG/HS seed-based connectivity (p_FDR_ = 0.018, r^2^_partial_ = 0.19); **Figure 3, Table S6**). HG/HS seed connectivity in right dorsal cerebellum (lobule VI) and orbitofrontal cortex were also modestly correlated with THI score, though below criteria for significance (p_FDR_<0.10, **Table S6**). These relationships were not present when measuring Yeo17 Dual Regression connectivity, or for other nodes tested in either model.

Follow-up analyses in SOC also identified modestly decreased auditory-network connectivity in people with chronic tinnitus using dual regression (e.g., r^2^_partial_ = 0.02, p_uncorr_ = 0.05; **Figure 3C**). Negative correlation between THI score and HG/HS seed network connectivity was also identified in right SOC (p_uncorr_=0.03, r^2^_partial_ [95% CI] = 0.06 [0.003,0.18]; **Figure 4D**).

**Figure 4.**
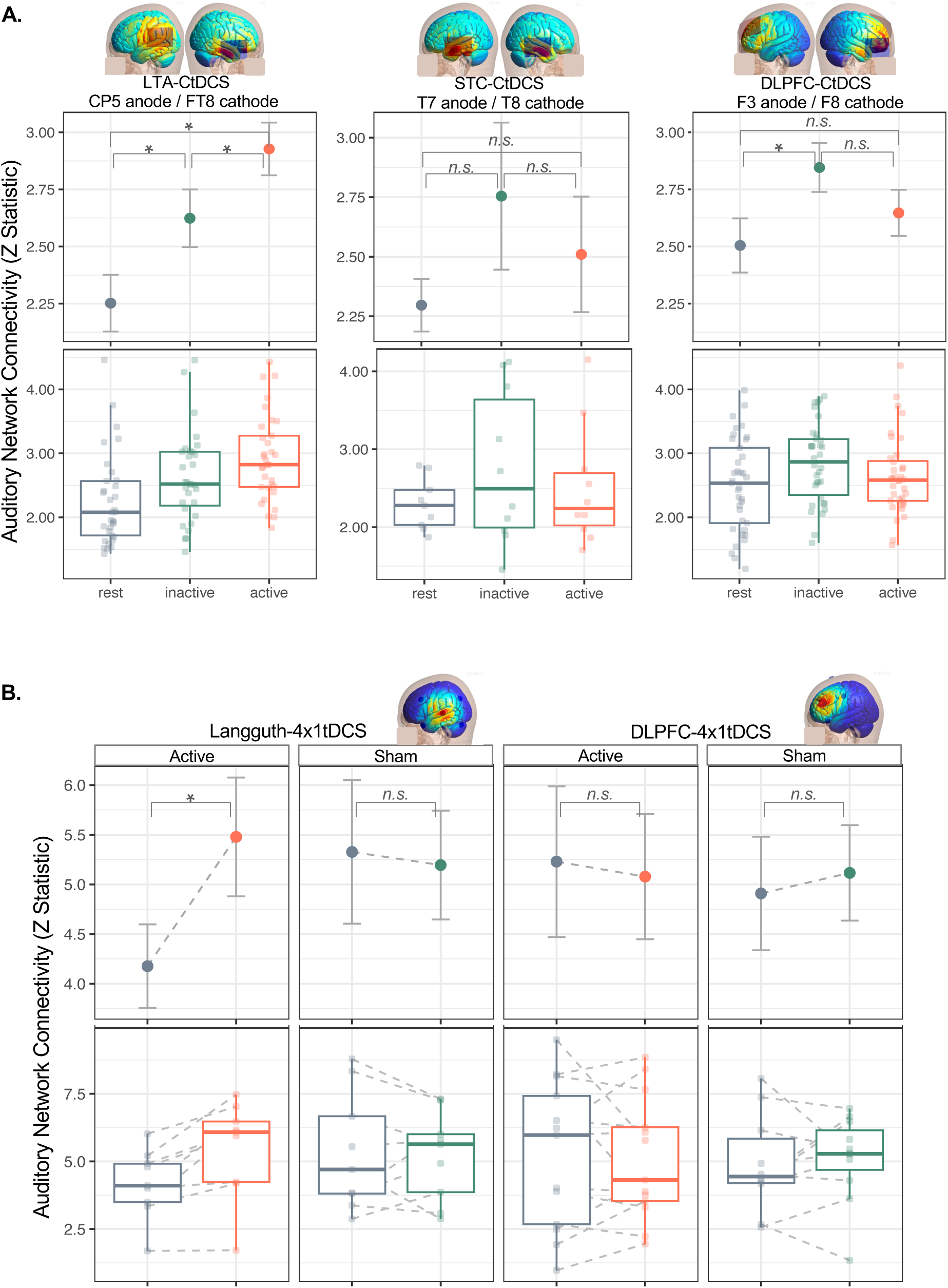
**A.** Auditory Network Strength increased during ∼5 minutes of active tDCS targeting left auditory cortex (LTA, left temporoparietal area) compared with inactive tDCS and rest. Network Strength did not differ during active and inactive STC- or DLPFC-tDCS. Top plots display mean and standard error for active, inactive, and rest conditions (orange, green, gray respectively), with corresponding boxplots below. Datapoints in boxplots reflect single scans. **B.** Auditory-network connectivity in right medial Heschl’s Gyrus (HG) increased after 20 minutes of active 4×1tDCS targeting left auditory cortex using the Langguth method. FC did not change in medial HG after inactive tDCS or active tDCS targeting left dorsolateral prefrontal cortex. Baseline (before tDCS) data are plotted in gray, with data after active and inactive tDCS in orange and green respectively. Data are otherwise plotted similar to A.

### Anodal tDCS Targeting Auditory Cortex Increases Auditory-Network Connectivity

In studies measuring the online effects of Conventional/Sponge tDCS (CtDCS), Auditory Network Strength increased during active LTA-CtDCS (**Figure 4; Table S7**). There was a significant main effect of tDCS condition (p_FDR_ < 0.05, F(2,61)=15.14), and Network Strength was highest during the active condition (active vs. inactive, t(61)=2.72, p_uncorr_<0.001; active vs. rest, t(60)=5.47, p_uncorr_<0.001; sham vs. rest, t(60)=2.73, p_uncorr_=0.008). Network Strength increased similarly in participants with and without chronic tinnitus during active LTA-C-tDCS; post hoc test for interaction between tDCS condition and tinnitus status was p > 0.05. Auditory Network Strength was not affected during active LTA-CtDCS when measuring Dual Regression connectivity, or during active STC- or DLPFC-CtDCS.

Nodewise effects did not reach statistical criterion for any CtDCS method when measuring connectivity using the HG/HS Seed or Dual Regression approach (**Tables S7-12**). However, some subthreshold effects are worth noting. HG/HS Seed connectivity modestly increased in left thalamus and left lateral HG during active LTA-CtDCS and DLPFC-CtDCS, respectively (both p_FDR_ < 0.10 for main effect of condition and active vs. inactive p_uncorr_ < 0.05; **Tables S7&9**). Auditory Network Strengh modestly decreased during active STC-CtDCS when measuring Dual Regresson connectivity (**Table S10**); note that in contrast to LTA-CtDCS, auditory cortex receives cathodal stimulation during STC-CtDCS (in addition to anodal stimulation, **Figure 1**).

In studies measuring the offline effects of 4×1tDCS on HG/HS Seed connectivity, all effects were p_FDR_ > 0.05 in both studies (**Tables S13&S14**). However, connectivity strength in the right medial HG node increased after active 4×1tDCS when targeting auditory cortex in tinnitus participants (condition-by-time interaction, t(16) = - 2.7, p_uncorr_ = 0.016; pre vs. post active tDCS, Cohen’s d = −1.6, p = 0.004; **Figure 4, Table S13**). When using the Dual Regression approach, additional HG nodes also showed modest increases after active 4×1tDCS targeting auditory cortex (p_uncorr_ < 0.05; **Tables S15**) similar to effects reported in the corresponding source study, which used dual regression [17]. These effects were not present after active 4×1tDCS targeting left DLPFC (**Table S16**).

## DISCUSSION

These retrospective analyses studied auditory-network connectivity and its modulation with tDCS in people with chronic tinnitus using pre-existing fMRI datasets [17, 22, 23]. Auditory network nodes were newly defined for this study, and included novel cerebellar nodes in addition to regions previously implicated in auditory-network connectivity and/or tinnitus (e.g., auditory cortex, prefrontal cortex, ventral striatum, etc. [47–49]). Corroborating recent evidence [7–10], auditory network strength was decreased in participants with chronic tinnitus, which could reflect increased spontaneous neuronal activity in the ascending auditory system as previously suggested [10, 11]. In a more novel finding, nodes in cerebellum and superior olivary complex were also less connected with the auditory network in chronic tinnitus, perhaps implicating a role for efferent cochlear pathways and/or head-centric somatosensory interference in chronic tinnitus [3, 50, 51]. Finally, single sessions of transcranial direct current stimulation increased auditory-network connectivity, laying the groundwork for potential “correction” of auditory-network discoherence in chronic tinnitus [17]. Taken together, this work sought to provide hypotheses and frameworks upon which future large-scale, balanced prospective studies of chronic tinnitus can be planned and executed.

### Auditory Network Discoherence in Chronic Tinnitus

In cross-sectional analyses comparing people with and without tinnitus, it is challenging (and perhaps impossible) to know whether differences identified are cause or consequence of the tinnitus. This limitation is not unique to tinnitus research, but is perhaps particularly insidious in this field given the disconnect between the simplicity of the tinnitus percept itself and the wide variety of systems it can impact (e.g., increased attentional load, annoyance, mental health, etc.). Even within the ascending auditory system, effects measured in one region could be caused by aberrant activity originating elsewhere within the system. Nevertheless, any consistent, measurable differences in chronic tinnitus participants that are independently validated across studies are potentially useful as biomarkers to facilitate diagnosis, assessment of treatment response, and/or identification of potential therapeutic targets.

The current study found reduced temporal coherence across the Auditory Network in people with chronic tinnitus measured with resting-state fMRI. Though a modest effect, it corroborates previous resting-state fMRI studies reporting reduced temporal coherence (i.e., functional connectivity) in the auditory system in people with tinnitus [7–10]. As previously suggested, this could relate to increased spontaneous neuronal activity reported in animal models of tinnitus [10, 11]. Empirical studies are needed to measure these relationships directly, for example to simultaneously measure spontaneous single-unit firing together with local field potentials in animal models. Given recent evidence that spontaneous firing in some neurons can covary with (i.e., is tuned to) specific local field potential frequencies, including low frequencies associated with BOLD fMRI [52], such frequency-minded analysis of spontaneous firing in animal models of tinnitus could be impactful.

If auditory-network discoherence continues to be identified across tinnitus datasets, it could be used as a biomarker in translational studies of treatment development. For example, tDCS protocols targeting auditory cortex with anodal stimulation in the current study increased auditory-network connectivity: more diffuse stimulation with conventional/sponge electrodes increased overall auditory-network connectivity and more focal stimulation with Ag/AgCl electrodes had more circumscribed effects in HG [17]. Notably, Network Strength decreased during STC-CtDCS where cathodal stimulation was applied to right auditory cortex (Figure 1), perhaps compatible with the idea that anodal stimulation increases neuronal excitability while cathodal stimulation decreases neuronal excitability [53]; however, these “trend-level” effects should be interpreted with caution given small sample size. If these tDCS protocols were improved to induce larger and/or long-lasting increases in auditory-network connectivity (e.g., increasing number of sessions, alternating current stimulation), this could lead to more consistent and durable effects on tinnitus symptoms. Such a biomarker could also be used to optimize settings for other neurostimulation (e.g., TMS, VNS) or acoustic stimulation studies, instead of or in addition to self-reported improvements. Though perhaps less useful as a stand-alone diagnostic biomarker given modest effect size in the current study, auditory-network discoherence may still contribute to diagnosis in combination with other items. Larger prospective studies with balanced hearing loss/history across groups are also needed to determine whether auditory-network discoherence could serve as a useful tinnitus biomarker.

### Effects in cerebellum and superior olivary complex (SOC) implicate head-centric interoception in tinnitus

Though the cerebellum is well-known to contribute to a variety of functions beyond its historically motor-centric definitions [54, 55], auditory-network definitions do not typically include the cerebellum. The current study identified auditory-network nodes in lobules VI and VIIIa and deep cerebellar nuclei, a novel contribution to auditory resting-state fMRI (see also [56]). In a related novel finding, participants with chronic tinnitus showed decreased auditory-network connectivity in these cerebellar nodes when controlling for their connectivity in other networks (i.e., in dual regression models). Participants with higher impact tinnitus also showed less connectivity between left lobule VI and the HG/HS seed-based auditory network, and complementary effects were also identified in the SOC. These results are similar to findings in idiopathic sudden sensorineural hearing loss and tinnitus, where functional connectivity between lobule VI and lateral auditory cortex (vs. medial HG/HS connectivity studied here) was also decreased [11]. Taken together, these results suggest discoherence in efferent cochlear pathways relevant to head-centric interoception in people with chronic tinnitus, particularly those with poorly compensated tinnitus. The remainder of this section discusses evidence in support of this idea.

Cerebellar lobule VI and VIIIa subregions identified in the current study contain somatomotor representations of articulators (e.g., mouth and tongue; [57, 58]), which may also explain their involvement in working memory, i.e., due to subvocalization [59, 60]. Lobule VI has also been linked with speech apraxia [61] and dysarthria [59, 62]. Notably, articulators are targeted in recent successful tinnitus trials via trigeminal nerve stimulation (i.e., jaw [50, 63] and tongue [64–66]), where paired auditory-somatosensory stimulation appears to be critical to efficacy [51, 65]. Thus, interactions between auditory-sensory and head-centric somatosensory systems clearly impact chronic tinnitus [3]. Indeed, tinnitus is typically perceived in a location inside the head or ear, and somatomotor manipulation of various parts of the head (e.g., eye, jaw) often impacts the tinnitus percept [67]. Head-centric interoception appears key.

Efferent cochlear pathways may also play a role in head-centric interoception. These pathways are well-known to modulate cochlear function (e.g., cochlear dynamics, cochlear nuclei activity) during loud noise and vocalization [68, 69]. Efferent olivocochlear pathways also change with age [70] and hearing loss [71], and have been implicated in chronic tinnitus in some human studies of otoacoustic emissions [72, 73] and animal models [74]. In the current study, SOC showed reduced connectivity in chronic tinnitus similar to cerebellar nodes in post hoc analyses. Future studies specifically designed to measure function in auditory brainstem structures [75] may be in a better position to measure SOC function, including delineation of its medial and lateral nuclei.

Cerebellar regions may interact with efferent cochlear pathways in different ways [76]. Though not identified by the current study, both the cerebellar vermis and paraflocculus receive auditory input, including from the SOC [77]. Early studies in guinea pigs identified efferent connections from the cerebellar vermis to olivary and cochlear nuclei [78], and reported that cerebellar stimulation decreased auditory nerve firing and cochlear microphonics, while cooling (inhibition) had the opposite effect [79]. In rodent models of tinnitus, the paraflocculus shows increased activity [80], and its modulation can reduce (or induce) behavioral and/or neuronal correlates of tinnitus in these animals [81–83]. It is unclear how these cerebellar regions with more established roles in auditory processing (paraflocculus, vermis) may interact with head-centric somatosensory regions (lobule VI, VIIa) identified in the current study. It is also worth noting that cerebellar regions may directly impact auditory cortex, inferior colliculus, and other auditory structures as well [74, 84]. More work is certainly needed, and there appears to be immense potential in studying these pathways to further mechanistic insight and advance treatment for chronic tinnitus [74, 82, 85].

### On the importance of defining auditory networks in resting-state fMRI

Resting-state fMRI studies are a powerful way of measuring network dynamics within the ascending auditory pathway; however, current standard approaches have limitations. In the popular Yeo17 atlas of resting-state networks for example, the auditory network is combined ventral somatomotor cortex [37]. It is unclear whether studies using this atlas are measuring auditory-network connectivity or a mixture of auditory and somatomotor network activity, thus potentially reducing their sensitivity to detect differences in nodes important to the auditory system. Yeo et al. acknowledged this, explaining that the combination of auditory and ventral somatomotor cortex in their data-driven approach could be due to smoothing and/or partial volume effects across the lateral fissure (i.e., between superior temporal and ventral somatomotor cortices; [37]).

The current study used an HG/HS seed-based approach to address this issue, where HG/HS seeds were defined using each participant’s superior temporal anatomy. Notably, this approach also identified two sets of auditory-network nodes in somatomotor cortex. The presence of somatomotor nodes in this HG/HS seed-based network is perhaps less likely to be due to partial volume effects across the lateral fissure, which raises the question of why these somatomotor nodes remain. Their involvement could be related to coordination of activity between auditory and somatomotor regions supporting vocalization, including own-voice sensory feedback [86, 87]. Though perhaps located too superior to overlap with primary representations of articulators (e.g., tongue), it is possible that these nodes overlap recently identified “intereffector” representations of complex movement where individual body parts appear not to be represented [88]. Independent validation of these results is needed, particularly in prospective studies better equipped to disentangle auditory-sensory vs. auditory-motor contributions in these nodes (e.g., including somatotopy). Here, the dual regression approach may be beneficial by controlling for primary somatosensory and other network signals when attempting to isolate auditory networks.

### Contributions of Hearing Loss and Depression to Reported Effects

People with chronic tinnitus often have elevated pure-tone thresholds and symptoms of depression [89]; therefore, controlling for potential contributions of depression status and mean pure-tone thresholds was an important aspect of the current statistical approach. Though not a primary focus of the current analyses, reduced Auditory Network Strength was noted in participants with depression, regardless of whether or not they had chronic tinnitus (**Table S3** and **Figure S1**). Thus, depression status may have effects on auditory-network connectivity independent of tinnitus, similar to previous reports of differences in sensory processing in depression perhaps related to anhedonia and/or attention [90, 91]. This underscores the importance of controlling for depressive symptoms and tinnitus-related distress in tinnitus research.

Hearing loss measured with pure-tone thresholds was not related to auditory network connectivity in these analyses. For example, overall Auditory Network Strength did not correlate with pure-tone thresholds (**Table S3**), and connectivity in single nodes also was not strongly correlated with hearing loss (data not shown). However, some studies have reported effects of hearing loss on functional connectivity specifically within auditory regions [92], and many more studies have identified effects outside auditory regions [93–97]. Though outside the scope of the current analyses, it is possible that averaging thresholds across all frequencies <=8kHz is too blunt an assessment when studying these functional changes, e.g. compared with isolating high frequency loss [93, 98]. Participants with different types and/or etiologies for their hearing loss could show also different functional changes within auditory networks (e.g., high frequency loss, [93]). A larger dataset where PTA thresholds and other aspects of hearing function are thoroughly assessed and properly balanced between tinnitus and controls (or studied independently of tinnitus) will be better able to address this issue.

### Conclusions

Though definitive causal mechanism(s) have yet to be established, converging evidence suggests chronic tinnitus is a problem of the brain and brain networks, not (only) of the ear. Many different brain regions and networks have been discussed as contributing and/or causative factors tinnitus, yet the common thread through all these is the tinnitus percept itself – typically a simple sound like high frequency tones or banded noise. The current study attempted to constrain analyses to auditory-sensory regions that are most likely to support perception of these simple sounds (e.g., HG/HS), and their highly connected neighbors in an Auditory Network. This approach corroborated evidence of decreased connectivity/coherence within the ascending auditory system [7–10] and presented novel fMRI evidence of decreased connectivity within cerebellar regions and efferent cochlear pathways – both relevant to head-centric interoception. It is still possible that regions outside the Auditory Network causally contribute to chronic tinnitus [12, 14, 99]; however, there are distinct advantages to identifying consistent tinnitus-related effects restricted to auditory-sensory regions and networks. Sampling the the entire brain using voxelwise analyses can be more prone to: (1) type I error due to the number of statistical tests (i.e., one test per ∼200,000 voxels in a liberal gray matter mask) and (2) the inclusion of distantly connected regions more likely to represent reactions to the tinnitus percept, and (perhaps) less relevant to its generation. There are limitations of the current study to consider, including that hearing loss was unbalanced across groups and not assessed in all participants, fMRI acquisition was not optimized for measuring brainstem function, and other aspects of tinnitus were not assessed (e.g., somatic contributions, sound level tolerance). Though the current approach to assessing functional connectivity was thorough, it is still possible that other approaches may yield different effects, e.g., using stricter temporal filtering [75, 100] or lagged/effective connectivity. Such alternative approaches may be more sensitive to detect tinnitus-related changes in inferior colliculus, for example, which were absent in the current study. Despite these limitations, the current study provides corroborating evidence for discoherence within auditory networks in chronic tinnitus (including efferent cochlear pathways), and lays groundwork for its correction with noninvasive brain stimulation. Future studies targeting interactions between head-centric auditory and somatomotor interoception may be particularly impactful.

## ACKNOWLEDGEMENTS

This work was supported by the NIH (R21DC015880), American Tinnitus Association, and NARSAD Young Investigator Grants from the Brain & Behavior Research Foundation (2016, 2020) to Dr. Leaver. This work was also supported in part through the computational resources and staff contributions provided for the Quest high performance computing facility at Northwestern University which is jointly supported by the Office of the Provost, the Office for Research, and Northwestern University Information Technology. This content is solely the responsibility of the author and does not necessarily reflect the official views of the National Institutes of Health or other sponsors.

## DECLARATIONS

The author declares that they have no competing interests. The author wrote and approved the final version of the manuscript being submitted, and warrants that the article is the author’s original work, has not received prior publication, and is not under consideration for publication elsewhere.

## Data Availability

Data is available from the author on reasonable request.

