## Supplement for "Auditory Network Discoherence in Chronic Tinnitus"

**Measuring and Modulating Auditory Network Discoherence in Chronic Tinnitus**

Amber M. Leaver Ph.D.[1]*
[1]Department of Radiology, Northwestern University, Chicago, IL, USA

### SUPPLEMENTAL METHODS

#### Participant Inclusion/Exclusion Criteria

For more detailed inclusion and exclusion criteria, refer to source publications.

Participants with chronic tinnitus reported having tinnitus for at least one year that was present when consciously attended for at least 10% of awake time for CtDCS studies and at least 50% of awake time for the 4x1tDCS study. Participants must have reported discussing tinnitus symptoms with a clinician to rule out obvious physical or neurological origin (e.g., acoustic neuroma, Meniere's Disease). Significant or severe developmental, neurological, or psychiatric conditions or substance abuse were also exclusion criteria, but mild-to-moderate mood or anxiety disorder and/or symptoms were not exclusionary for tinnitus studies.

Participants with depression reported having a diagnosis of major depression at least one year prior to participating in the study, with mild-to-moderate symptoms. As for all participants, acute suicidality was exclusionary, as were other neurological, developmental, or other health conditions affecting brain function. Comorbid diagnosis or symptoms of anxiety were not exclusionary for participants with depression.

For all participants, any change in treatment within 6 weeks of study start was exclusionary (e.g., SSRIs). Participants having had brain stimulation within 6 weeks of study start were also excluded.

#### MRI Acquisition

MR images were acquired using 3T Prisma scanners at the UCLA Brain Mapping Center and Northwestern (NU) Center for Translational Imaging using identical sequences. Sequence parameters for BOLD-fMRI scans were as follows: 2mm isotropic, 0.8s repetition time (TR), 37ms echo time (TE), 52° flip angle, 72 axial slices, 8 multiband factor. T1- and T2-weighted anatomical scans were also acquired: T1 multi-echo MPRAGE 0.8mm isotropic, TR=2.5s, TE=1.8, 3.6, 5.4, 7.2ms combined, 1000ms inversion time, 8 degree flip angle; T2 SPACE 0.8mm isotropic, TR=3.2s, TE=564ms (effective TE 559.7ms), echo train length = 1166ms.

#### Defining the Auditory Network (AudNet)

As described in the main text, auditory-network nodes were defined using voxelwise HG/HS seed maps averaged across all participants (MNI space). This average map was thresholded at mean Z values ranging from 1.33 to 4.67 (approximately $p < 0.10$ to 0.000001) and visually inspected to select a threshold for each region that balanced cluster size with corresponding region.

It was expected that functional connectivity values would be high in auditory cortex near the HG/HS seed regions, and lower in more distantly connected and difficult-to-image regions. Therefore, considering a range of thresholds allowed a more inclusive approach to node definition, e.g., including regions like parahippocampal gyrus and medial geniculate nucleus that were present at lower thresholds.

Note that in this approach, an average value of $z > 1.33$ ($p < 0.10$) does not indicate an absence of statistically "significant" connectivity, but rather an average value across all participants and scans. Follow-up testing confirmed that connectivity was above zero using one-sample t tests $p_{FDR} < 0.05$.

#### Transcranial Direct Current Stimulation (tDCS)

Transcranial direct current stimulation (tDCS) was delivered using a Soterix 1x1 tES device and 4x1 HD-tDCS conversion box. Current was constant for active tDCS with 30 second linear on- and off-ramps. Current was ramped up and down at the beginning and end of the session for sham stimulation (as programmed by the manufacturer). Electrodes were positioned with reference to the 10-10 EEG system as described in the main text (**Figure 1**) and source publications.

For Conventional tDCS (CtDCS),[2] stimulation occurred during BOLD fMRI scans to measure online effects of tDCS on resting-state functional connectivity. Current amplitude was 2mA (i.e., +2mA anode and -2mA cathode) and stimulation was ~5 minutes in duration. Two 5x7cm$^2$ sponges wetted with saline (~7mL per electrode) were each fitted to a 5x5mm$^2$ carbon rubber electrode. A thin layer of conductive paste and gel were also applied to the rubber electrode and sponge electrode cover, respectively, to prevent drying during the hour-long MRI scan. Electrodes were positioned on the volunteer's head immediately before the MRI and secured with broad flexible bands made of rubber or vinyl. Electrode positions were checked visually before and after the scan, and during the scan by noting the position of the electrode on T1- and T2-weighted anatomical images.

For 4x1 tDCS studies, offline effects were measured, comparing BOLD-fMRI scans before and after stimulation. Current amplitude was 2mA (i.e., +2mA anode and -0.5mA each cathode) and stimulation was 20 minutes in duration. Small Ag/AgCl electrodes were placed in a 4x1 configuration (i.e., High Definition or HD, *Soterix*) with center anode and surround cathodes positioned as described in main text. Electrodes were positioned in custom plastic holders affixed to a spandex cap and filled with Lectron II conductive gel (~1.5mL/electrode).

**Statistics: Follow-up analysis of depression and chronic tinnitus**

Though the focus of this manuscript was chronic tinnitus, we report a follow-up analysis of potential interactions between tinnitus and depression status on Auditory Network Strength measured with HG/HS seed-based connectivity and using the complete dataset. Tinnitus and depression were factors of interest, and age, sex, site, mean relative motion, session, and tDCS were fixed factors of no interest, and subject was a factor of no interest, just as in the main analyses. Main effects of tinnitus and depression are reported along with statistics for an interaction between tinnitus and depression (**Figure S1**).

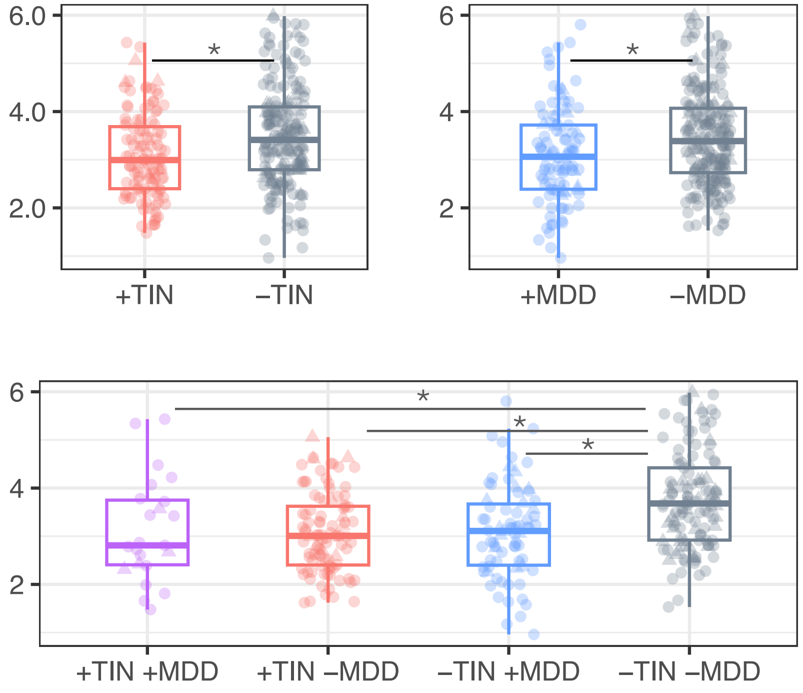


**Figure S1.** Auditory Network Strength was reduced in people with chronic tinnitus (+TIN) and depression (+MDD) compared to controls (-TIN, -MDD). **A-B.** Box plots display data for people with and without chronic tinnitus and depression, and asterisks mark significant main effects of tinnitus (p=0.012) and depression (p=0.002) on Network Strength measured using HG/HS Seed connectivity and all data. **C.** An interaction between depression and tinnitus groups was not significant (t(102)=1.95,p=0.054). Although Network Strength is less in each group compared with the control group (-TIN –MDD; p< 0.03 for all), participants with both chronic and tinnitus and depression (+TIN+MDD) were no different from tinnitus only (+TIN-MDD) or depression only (-TIN+MDD) groups (p>0.90 for both).

**Table S1. Participant Characteristics by Tinnitus and Depression Status**

| | +TIN +MDD | +TIN -MDD | -TIN +MDD | -TIN -MDD | Statistical Comparisons |
|---|---|---|---|---|---|
| Sample Size | 8 | 35 | 27 | 47 | NR |
| Site, NU (UCLA) | 6 (2) | 29 (6) | 19 (8) | 30 (17) | *X^2(7)=57.59, p=4.56E-10* |
| Sex, F (M) | 3 (5) | 18 (17) | 15 (12) | 26 (21) | *X^2(7)=28.98, p=0.0001* |
| Age, mean (SD) in yrs | 45.25 (12.23) | 51.60 (13.24) | 30.63 (9.63) | 33.53 (13.35) | *F(3,113)=19.59, p=2.69E-10* |
| PT Threshold, mean (SD) in dB | 22.92 (13.34) | 27.26 (18.11) | 12.94 (2.33) | 10.80 (5.25) | *F(3,72)=8.42, p=0.00007* |
| Tinnitus Handicap Inventory, mean (SD) | 52.86 (16.51) | 49.53 (17.81) | NA | NA | *Welch's t(9)=0.47, p=0.65* |
| Tinnitus Functional Index, mean (SD) | 30.72 (27.92) | 38.55 (21.82) | NA | NA | *Welch's t(5)=-0.59, p=0.58* |
| Beck Depression Inventory, mean (SD) | 11.29 (6.92) | 6.87 (6.80) | 25.24 (10.06) | 3.39 (5.45) | *F(3,108)=52.64, p=4.77E-21* |
| Mean Rel. Motion, mean (SD) in mm | 0.14 (0.05) | 0.10 (0.04) | 0.13 (0.06) | 0.11 (0.06) | *F(3,113)=1.68, p=0.18* |

*Statistical tests are Chi Squared, One-way ANOVA, or Welch's t test (unbalanced sample size).*

**Table S2. Auditory Network Nodes**

| Node Label | Full Name | Definition Method | Center of Gravity $X_{MNI}, Y_{MNI}, Z_{MNI}$ |
|---|---|---|---|
| R_aIns | Right anterior insula | z < 4.67, Group Average HG/HS Seed Map | 37, 9.04, 8 |
| L_aIns | Left anterior insula | z < 4.33, Group Average HG/HS Seed Map | -35.8, 5.12, 9 |
| R_CS | Right central sulcus | z < 4.00, Group Average HG/HS Seed Map | 48.8, -10.3, 41.8 |
| L_CS | Left central sulcus | z < 4.00, Group Average HG/HS Seed Map | -41.8, -16.6, 42 |
| preSMA | Presupplementary Motor Area | z < 3.67, Group Average HG/HS Seed Map | 0.301, 10.6, 39.3 |
| R_dCS | Right dorsal central sulcus | z < 3.33, Group Average HG/HS Seed Map | 21.5, -27.8, 60.4 |
| L_dCS | Left dorsal central sulcus | z < 3.33, Group Average HG/HS Seed Map | -19.8, -28.6, 61.1 |
| rACC | Right anterior cingulate cortex | z < 3.33, Group Average HG/HS Seed Map | 1.19, 38.4, 12.4 |
| L_Th | Left dorsal thalamus | z < 2.67, Group Average HG/HS Seed Map | -8.12, -17.8, 6.76 |
| R_Th | Right dorsal thalamus | z < 2.67, Group Average HG/HS Seed Map | 9.39, -16.1, 7.54 |
| L_MGN | Left medial geniculate nucleus | z < 1.67, Group Average HG/HS Seed Map | -12.6, -28.4, -4.4 |
| R_MGN | Right medial geniculate nucleus | z < 1.67, Group Average HG/HS Seed Map | 13.1, -28.1, -3.71 |
| R_DLPFC | Right dorsolateral prefrontal cortex | z < 1.67, Group Average HG/HS Seed Map | 31.1, 43.7, 23.3 |
| R_OFC | Right orbitofrontal cortex | z < 1.67, Group Average HG/HS Seed Map | 25.2, 31.4, -14.5 |
| L_OFC | Left orbitofrontal cortex | z < 1.67, Group Average HG/HS Seed Map | -26.9, 34.8, -13.1 |
| R_vCB | Right ventral cerebellum | z < 1.67, Group Average HG/HS Seed Map | 13.7, -67.2, -49 |
| L_vCB | Left ventral cerebellum | z < 1.67, Group Average HG/HS Seed Map | -16.1, -66.1, -51.1 |
| L_DLPFC | Left dorsolateral prefrontal cortex | z < 1.33, Group Average HG/HS Seed Map | -32.3, 41.9, 2 |
| R_dCB | Right dorsal cerebellum | z < 1.33, Group Average HG/HS Seed Map | 16.5, -61.6, -19.5 |
| L_dCB | Left dorsal cerebellum | z < 1.33, Group Average HG/HS Seed Map | -15.4, -62.5, -19.4 |
| R_DN | Right deep nucleus, cerebellum | z < 1.33, Group Average HG/HS Seed Map | 14 -54.2 -34.2 |
| L_DN | Left deep nucleus, cerebellum | z < 1.33, Group Average HG/HS Seed Map | -14.2 -53.8 -34.7 |
| R_PHG | Right parahippocampal cortex | z < 1.33, Group Average HG/HS Seed Map | 29.5, -12.8, -31 |
| L_PHG | Left parahippocampal cortex | z < 1.33, Group Average HG/HS Seed Map | -28.6, -12.3, -33.7 |
| R_IFG | Right inferior frontal gyrus | z < 1.33, Group Average HG/HS Seed Map | 41.7, 41.2, 7.46 |
| L_VS | Left ventral striatum | z < 2.33, Group Average HG/HS Seed Map | -22.5, 4.96, -9.93 |
| R_VS | Right ventral striatum | z < 2.33, Group Average HG/HS Seed Map | 22, 6.62, -11 |
| L_VC | Left medial visual cortex | z < 2.33, Group Average HG/HS Seed Map | -14.1, -68.8, 7.19 |
| R_VC | Right medial visual cortex | z < 2.33, Group Average HG/HS Seed Map | 20.1, -61.4, 6.63 |
| L_HG | Left Heschl's Gyrus | Jülich Atlas TE1.0 | N/A |
| R_HG | Right Heschl's Gyrus | Jülich Atlas TE1.0 | N/A |
| L_mHG | Left medial Heschl's Gyrus | Jülich Atlas TE1.1 | N/A |
| R_mHG | Right medial Heschl's Gyrus | Jülich Atlas TE1.1 | N/A |
| L_lHG | Left lateral Heschl's Gyrus | Jülich Atlas TE1.2 | N/A |
| R_lHG | Right lateral Heschl's Gyrus | Jülich Atlas TE1.2 | N/A |
| L_IC | Left Inferior colliculus | ID'ed using anatomical features on MNI Template | -4, -36, -12 |
| R_IC | Right inferior colliculus | ID'ed using anatomical features on MNI Template | 6,-36, -12 |

**Table S3. Auditory Network Strength Across Model Terms**

| | Model Term | r2 | r2_upperCI | r2_lowerCI | p |
|---|---|---|---|---|---|
| Complete Dataset | Full Model | 0.2688 | 0.3619 | 0.2070 | <0.0001 |
| | tinnitus | 0.0433 | 0.0970 | 0.0100 | 0.0117 |
| | age | 0.0227 | 0.0662 | 0.0017 | 0.0702 |
| | sex | 0.0073 | 0.0381 | 0.0000 | 0.2930 |
| | depression | 0.0619 | 0.1220 | 0.0205 | 0.0023 |
| | site | 0.0420 | 0.0952 | 0.0094 | 0.0038 |
| | rel_mean | 0.0286 | 0.0755 | 0.0035 | 0.0193 |
| | session | 0.0656 | 0.1267 | 0.0227 | <0.0001 |
| | task_inactive | 0.0949 | 0.1657 | 0.0449 | 0.4468 |
| | task_rest | NA | NA | NA | <0.0001 |
| Partial, PTA | Full Model | 0.2235 | 0.3391 | 0.1627 | <0.0001 |
| | tinnitus | 0.0325 | 0.0915 | 0.0028 | 0.0839 |
| | mean PTA | 0.0131 | 0.0578 | 0.0001 | 0.2654 |
| | age | 0.0289 | 0.0858 | 0.0018 | 0.1186 |
| | sex | 0.0123 | 0.0563 | 0.0001 | 0.2983 |
| | depression | 0.0188 | 0.0688 | 0.0003 | 0.2101 |
| | site | 0.0110 | 0.0537 | 0.0001 | 0.2320 |
| | rel_mean | 0.0484 | 0.1151 | 0.0089 | 0.0139 |
| | session | 0.0890 | 0.1683 | 0.0317 | <0.0001 |
| | task_inactive | 0.0678 | 0.1457 | 0.0219 | 0.5699 |
| | task_rest | NA | NA | NA | 0.0006 |
| Partial, Balanced PTA | Full Model | 0.3150 | 0.4359 | 0.2407 | <0.0001 |
| | tinnitus | 0.0484 | 0.1225 | 0.0067 | 0.0449 |
| | mean PTA | 0.0069 | 0.0496 | 0.0000 | 0.4587 |
| | age | 0.0456 | 0.1186 | 0.0057 | 0.0585 |
| | sex | 0.0194 | 0.0760 | 0.0002 | 0.2096 |
| | depression | 0.0829 | 0.1691 | 0.0241 | 0.0130 |
| | site | 0.0097 | 0.0564 | 0.0000 | 0.2856 |
| | rel_mean | 0.0477 | 0.1216 | 0.0065 | 0.0252 |
| | session | 0.1089 | 0.2011 | 0.0405 | <0.0001 |
| | task_inactive | 0.0531 | 0.1346 | 0.0120 | 0.4955 |
| | task_rest | NA | NA | NA | 0.0111 |

**Table S4. Nodewise HG/HS Seed Auditory Network Connectivity in Chronic Tinnitus**

| Model Description | roi | t | df | r2 | r2_uCI | r2_lCI | p | pfdr |
|---|---|---|---|---|---|---|---|---|
| Complete | L_HG | 1.569 | 99.829 | 0.019 | 0.061 | 0.001 | 0.120 | 0.554 |
| Partial, PTA | L_HG | 1.891 | 66.772 | 0.041 | 0.105 | 0.006 | 0.063 | 0.545 |
| Partial, PTA<30dB | L_HG | 2.141 | 55.534 | 0.054 | 0.131 | 0.009 | 0.037 | 0.339 |
| Complete | R_HG | 2.064 | 97.040 | 0.032 | 0.081 | 0.005 | 0.042 | 0.361 |
| Partial, PTA | R_HG | 2.293 | 62.436 | 0.053 | 0.121 | 0.011 | 0.025 | 0.369 |
| Partial, PTA<30dB | R_HG | 2.579 | 52.194 | 0.075 | 0.159 | 0.019 | 0.013 | 0.266 |
| Complete | L_lHG | 0.896 | 99.296 | 0.006 | 0.035 | 0.000 | 0.373 | 0.733 |
| Partial, PTA | L_lHG | 0.669 | 65.102 | 0.005 | 0.039 | 0.000 | 0.506 | 0.921 |
| Partial, PTA<30dB | L_lHG | 0.523 | 52.707 | 0.004 | 0.041 | 0.000 | 0.603 | 0.892 |
| Complete | R_lHG | 1.951 | 103.867 | 0.029 | 0.076 | 0.004 | 0.054 | 0.361 |
| Partial, PTA | R_lHG | 2.238 | 69.090 | 0.056 | 0.125 | 0.013 | 0.028 | 0.369 |
| Partial, PTA<30dB | R_lHG | 2.527 | 55.518 | 0.084 | 0.171 | 0.025 | 0.014 | 0.266 |
| Complete | L_mHG | 2.010 | 100.329 | 0.032 | 0.081 | 0.005 | 0.047 | 0.361 |
| Partial, PTA | L_mHG | 2.219 | 66.312 | 0.057 | 0.127 | 0.013 | 0.030 | 0.369 |
| Partial, PTA<30dB | L_mHG | 2.333 | 54.652 | 0.070 | 0.152 | 0.017 | 0.023 | 0.288 |
| Complete | R_mHG | 1.204 | 100.778 | 0.012 | 0.047 | 0.000 | 0.231 | 0.686 |
| Partial, PTA | R_mHG | 0.688 | 64.788 | 0.005 | 0.041 | 0.000 | 0.494 | 0.921 |
| Partial, PTA<30dB | R_mHG | 0.567 | 53.698 | 0.004 | 0.043 | 0.000 | 0.573 | 0.887 |
| Complete | L_MGN | 0.635 | 94.650 | 0.002 | 0.025 | 0.000 | 0.527 | 0.733 |
| Partial, PTA | L_MGN | 0.530 | 67.059 | 0.002 | 0.030 | 0.000 | 0.598 | 0.921 |
| Partial, PTA<30dB | L_MGN | 0.564 | 53.658 | 0.003 | 0.037 | 0.000 | 0.575 | 0.887 |
| Complete | R_MGN | 1.215 | 99.579 | 0.009 | 0.043 | 0.000 | 0.227 | 0.686 |
| Partial, PTA | R_MGN | 1.453 | 67.837 | 0.017 | 0.066 | 0.000 | 0.151 | 0.697 |
| Partial, PTA<30dB | R_MGN | 0.980 | 53.876 | 0.010 | 0.057 | 0.000 | 0.331 | 0.843 |
| Complete | L_IC | 0.593 | 99.880 | 0.002 | 0.024 | 0.000 | 0.555 | 0.733 |
| Partial, PTA | L_IC | 0.049 | 66.169 | 0.000 | 0.022 | 0.000 | 0.961 | 0.988 |
| Partial, PTA<30dB | L_IC | -0.077 | 54.316 | 0.000 | 0.027 | 0.000 | 0.939 | 0.957 |
| Complete | R_IC | 0.776 | 90.746 | 0.003 | 0.028 | 0.000 | 0.440 | 0.733 |
| Partial, PTA | R_IC | 0.252 | 62.319 | 0.000 | 0.024 | 0.000 | 0.801 | 0.988 |
| Partial, PTA<30dB | R_IC | 0.339 | 53.546 | 0.001 | 0.031 | 0.000 | 0.736 | 0.937 |
| Complete | L_aIns | 0.319 | 105.560 | 0.001 | 0.019 | 0.000 | 0.750 | 0.868 |
| Partial, PTA | L_aIns | 0.592 | 67.081 | 0.004 | 0.036 | 0.000 | 0.556 | 0.921 |
| Partial, PTA<30dB | L_aIns | 0.764 | 56.313 | 0.007 | 0.050 | 0.000 | 0.448 | 0.843 |
| Complete | L_CS | 1.913 | 101.224 | 0.020 | 0.063 | 0.001 | 0.059 | 0.361 |
| Partial, PTA | L_CS | 1.483 | 69.290 | 0.017 | 0.066 | 0.000 | 0.143 | 0.697 |
| Partial, PTA<30dB | L_CS | 1.484 | 55.790 | 0.023 | 0.084 | 0.000 | 0.143 | 0.663 |
| Complete | L_dCB | 0.165 | 96.095 | 0.000 | 0.017 | 0.000 | 0.869 | 0.946 |
| Partial, PTA | L_dCB | -0.172 | 66.513 | 0.000 | 0.023 | 0.000 | 0.864 | 0.988 |
| Partial, PTA<30dB | L_dCB | -0.282 | 52.063 | 0.001 | 0.031 | 0.000 | 0.779 | 0.937 |
| Complete | L_dCS | 2.082 | 97.045 | 0.023 | 0.067 | 0.002 | 0.040 | 0.361 |
| Partial, PTA | L_dCS | 1.102 | 68.212 | 0.009 | 0.050 | 0.000 | 0.274 | 0.867 |
| Partial, PTA<30dB | L_dCS | 1.253 | 54.433 | 0.016 | 0.069 | 0.000 | 0.215 | 0.725 |
| Complete | L_DLPFC | -0.068 | 102.975 | 0.000 | 0.016 | 0.000 | 0.946 | 0.972 |
| Partial, PTA | L_DLPFC | 0.220 | 67.755 | 0.000 | 0.024 | 0.000 | 0.827 | 0.988 |
| Partial, PTA<30dB | L_DLPFC | 0.293 | 53.797 | 0.001 | 0.032 | 0.000 | 0.770 | 0.937 |
| Complete | L_DN | 1.050 | 89.931 | 0.005 | 0.034 | 0.000 | 0.297 | 0.686 |
| Partial, PTA | L_DN | 0.902 | 61.110 | 0.006 | 0.041 | 0.000 | 0.371 | 0.921 |
| Partial, PTA<30dB | L_DN | 0.861 | 51.228 | 0.007 | 0.049 | 0.000 | 0.393 | 0.843 |
| Complete | L_OFC | -0.798 | 97.975 | 0.005 | 0.032 | 0.000 | 0.427 | 0.733 |
| Partial, PTA | L_OFC | -1.819 | 63.751 | 0.031 | 0.089 | 0.002 | 0.074 | 0.545 |
| Partial, PTA<30dB | L_OFC | -1.609 | 49.721 | 0.025 | 0.086 | 0.001 | 0.114 | 0.602 |
| Complete | L_PHG | 2.068 | 96.221 | 0.023 | 0.067 | 0.002 | 0.041 | 0.361 |
| Partial, PTA | L_PHG | 1.528 | 64.708 | 0.017 | 0.066 | 0.000 | 0.131 | 0.697 |
| Partial, PTA<30dB | L_PHG | 1.846 | 51.653 | 0.028 | 0.091 | 0.001 | 0.071 | 0.523 |
| Complete | L_Th | 0.615 | 97.367 | 0.002 | 0.026 | 0.000 | 0.540 | 0.733 |
| Partial, PTA | L_Th | 0.270 | 65.376 | 0.001 | 0.025 | 0.000 | 0.788 | 0.988 |
| Partial, PTA<30dB | L_Th | 0.188 | 51.499 | 0.000 | 0.028 | 0.000 | 0.851 | 0.937 |
| Complete | L_VC | -1.092 | 90.343 | 0.007 | 0.037 | 0.000 | 0.278 | 0.686 |
| Partial, PTA | L_VC | 0.146 | 62.244 | 0.000 | 0.023 | 0.000 | 0.884 | 0.988 |
| Partial, PTA<30dB | L_VC | -0.055 | 48.646 | 0.000 | 0.027 | 0.000 | 0.957 | 0.957 |
| Complete | L_vCB | 0.187 | 102.567 | 0.000 | 0.018 | 0.000 | 0.852 | 0.946 |
| Partial, PTA | L_vCB | -0.208 | 67.502 | 0.000 | 0.024 | 0.000 | 0.836 | 0.988 |

| | | | | | | | | |
|---|---|---|---|---|---|---|---|---|
| Partial, PTA<30dB | L_vCB | -0.254 | 53.519 | 0.001 | 0.031 | 0.000 | 0.801 | 0.937 |
| Complete | L_VS | 1.121 | 95.662 | 0.008 | 0.040 | 0.000 | 0.265 | 0.686 |
| Partial, PTA | L_VS | 1.035 | 65.833 | 0.009 | 0.049 | 0.000 | 0.305 | 0.867 |
| Partial, PTA<30dB | L_VS | 1.269 | 53.806 | 0.015 | 0.067 | 0.000 | 0.210 | 0.725 |
| Complete | preSMA | 1.439 | 98.990 | 0.013 | 0.049 | 0.000 | 0.153 | 0.630 |
| Partial, PTA | preSMA | 0.775 | 68.360 | 0.005 | 0.040 | 0.000 | 0.441 | 0.921 |
| Partial, PTA<30dB | preSMA | 0.801 | 53.366 | 0.006 | 0.048 | 0.000 | 0.427 | 0.843 |
| Complete | R_aIns | 1.203 | 99.804 | 0.009 | 0.042 | 0.000 | 0.232 | 0.686 |
| Partial, PTA | R_aIns | 1.387 | 63.839 | 0.018 | 0.067 | 0.000 | 0.170 | 0.699 |
| Partial, PTA<30dB | R_aIns | 1.705 | 56.398 | 0.030 | 0.094 | 0.001 | 0.094 | 0.578 |
| Complete | R_CS | 0.889 | 99.110 | 0.005 | 0.033 | 0.000 | 0.376 | 0.733 |
| Partial, PTA | R_CS | 1.163 | 65.122 | 0.012 | 0.056 | 0.000 | 0.249 | 0.867 |
| Partial, PTA<30dB | R_CS | 1.257 | 53.364 | 0.017 | 0.073 | 0.000 | 0.214 | 0.725 |
| Complete | R_dCB | 0.629 | 103.147 | 0.003 | 0.027 | 0.000 | 0.531 | 0.733 |
| Partial, PTA | R_dCB | 0.462 | 69.581 | 0.002 | 0.031 | 0.000 | 0.646 | 0.956 |
| Partial, PTA<30dB | R_dCB | 0.283 | 54.117 | 0.001 | 0.031 | 0.000 | 0.778 | 0.937 |
| Complete | R_dCS | 1.072 | 98.846 | 0.007 | 0.037 | 0.000 | 0.286 | 0.686 |
| Partial, PTA | R_dCS | 0.551 | 66.692 | 0.002 | 0.032 | 0.000 | 0.584 | 0.921 |
| Partial, PTA<30dB | R_dCS | 0.764 | 55.039 | 0.006 | 0.047 | 0.000 | 0.448 | 0.843 |
| Complete | R_DLPFC | 0.369 | 97.163 | 0.001 | 0.020 | 0.000 | 0.713 | 0.851 |
| Partial, PTA | R_DLPFC | 0.553 | 64.354 | 0.003 | 0.033 | 0.000 | 0.582 | 0.921 |
| Partial, PTA<30dB | R_DLPFC | 0.810 | 50.642 | 0.007 | 0.050 | 0.000 | 0.422 | 0.843 |
| Complete | R_DN | 0.875 | 98.393 | 0.004 | 0.031 | 0.000 | 0.384 | 0.733 |
| Partial, PTA | R_DN | 1.065 | 67.257 | 0.009 | 0.049 | 0.000 | 0.291 | 0.867 |
| Partial, PTA<30dB | R_DN | 0.800 | 54.865 | 0.006 | 0.048 | 0.000 | 0.427 | 0.843 |
| Complete | R_IFG | 1.597 | 94.870 | 0.015 | 0.054 | 0.000 | 0.113 | 0.554 |
| Partial, PTA | R_IFG | -0.115 | 62.536 | 0.000 | 0.023 | 0.000 | 0.909 | 0.988 |
| Partial, PTA<30dB | R_IFG | 0.176 | 49.420 | 0.000 | 0.028 | 0.000 | 0.861 | 0.937 |
| Complete | R_OFC | 0.106 | 93.528 | 0.000 | 0.017 | 0.000 | 0.916 | 0.968 |
| Partial, PTA | R_OFC | 0.014 | 62.092 | 0.000 | 0.022 | 0.000 | 0.989 | 0.989 |
| Partial, PTA<30dB | R_OFC | 0.298 | 49.699 | 0.001 | 0.031 | 0.000 | 0.767 | 0.937 |
| Complete | R_PHG | 0.452 | 98.272 | 0.001 | 0.022 | 0.000 | 0.653 | 0.805 |
| Partial, PTA | R_PHG | 0.570 | 65.892 | 0.003 | 0.033 | 0.000 | 0.570 | 0.921 |
| Partial, PTA<30dB | R_PHG | 0.866 | 54.229 | 0.007 | 0.052 | 0.000 | 0.390 | 0.843 |
| Complete | R_Th | 0.505 | 93.985 | 0.002 | 0.023 | 0.000 | 0.614 | 0.784 |
| Partial, PTA | R_Th | -0.049 | 62.718 | 0.000 | 0.022 | 0.000 | 0.961 | 0.988 |
| Partial, PTA<30dB | R_Th | -0.232 | 50.829 | 0.001 | 0.029 | 0.000 | 0.817 | 0.937 |
| Complete | R_VC | -0.034 | 95.508 | 0.000 | 0.016 | 0.000 | 0.973 | 0.973 |
| Partial, PTA | R_VC | 0.316 | 64.158 | 0.001 | 0.026 | 0.000 | 0.753 | 0.988 |
| Partial, PTA<30dB | R_VC | 0.058 | 50.433 | 0.000 | 0.027 | 0.000 | 0.954 | 0.957 |
| Complete | R_vCB | 0.761 | 100.756 | 0.004 | 0.029 | 0.000 | 0.448 | 0.733 |
| Partial, PTA | R_vCB | 0.875 | 67.732 | 0.007 | 0.044 | 0.000 | 0.385 | 0.921 |
| Partial, PTA<30dB | R_vCB | 0.714 | 53.370 | 0.005 | 0.046 | 0.000 | 0.478 | 0.843 |
| Complete | R_VS | 0.739 | 93.876 | 0.003 | 0.028 | 0.000 | 0.462 | 0.733 |
| Partial, PTA | R_VS | 0.371 | 64.939 | 0.001 | 0.027 | 0.000 | 0.712 | 0.988 |
| Partial, PTA<30dB | R_VS | 0.718 | 54.252 | 0.004 | 0.043 | 0.000 | 0.476 | 0.843 |
| Complete | rACC | 0.709 | 101.797 | 0.003 | 0.026 | 0.000 | 0.480 | 0.733 |
| Partial, PTA | rACC | 0.547 | 74.316 | 0.002 | 0.031 | 0.000 | 0.586 | 0.921 |
| Partial, PTA<30dB | rACC | 0.642 | 62.070 | 0.003 | 0.040 | 0.000 | 0.523 | 0.880 |

**Nodes pFDR < 0.05 and nodes [t]pFDR < 0.10 are marked in bold italic font.*

**Table S5. Nodewise Yeo17 (Dual Regression) Auditory Network Connectivity in Chronic Tinnitus**

| Model Description | roi | t | df | r2 | r2_uCI | r2_lCI | p | $p_{FDR}$ |
|---|---|---|---|---|---|---|---|---|
| Complete | L_HG | 0.369 | 96.095 | 0.001 | 0.021 | 0.000 | 0.713 | 0.859 |
| Partial, PTA | L_HG | 1.172 | 66.889 | 0.013 | 0.057 | 0.000 | 0.245 | 0.740 |
| Partial, PTA<30dB | L_HG | 1.294 | 54.370 | 0.017 | 0.071 | 0.000 | 0.201 | 0.561 |
| Complete | R_HG | 1.387 | 100.703 | 0.015 | 0.052 | 0.000 | 0.169 | 0.438 |
| Partial, PTA | R_HG | 1.210 | 67.062 | 0.016 | 0.062 | 0.000 | 0.231 | 0.740 |
| Partial, PTA<30dB | R_HG | 1.409 | 53.447 | 0.025 | 0.085 | 0.001 | 0.164 | 0.561 |
| Complete | L_lHG | -0.008 | 94.687 | 0.000 | 0.016 | 0.000 | 0.993 | 0.993 |
| Partial, PTA | L_lHG | 0.104 | 67.739 | 0.000 | 0.022 | 0.000 | 0.917 | 0.933 |
| Partial, PTA<30dB | L_lHG | -0.157 | 60.105 | 0.000 | 0.027 | 0.000 | 0.876 | 0.929 |
| Complete | R_lHG | 0.188 | 96.318 | 0.000 | 0.017 | 0.000 | 0.851 | 0.900 |
| Partial, PTA | R_lHG | -0.113 | 66.865 | 0.000 | 0.022 | 0.000 | 0.911 | 0.933 |
| Partial, PTA<30dB | R_lHG | 0.092 | 53.031 | 0.000 | 0.026 | 0.000 | 0.927 | 0.953 |
| Complete | L_mHG | 1.120 | 92.153 | 0.009 | 0.040 | 0.000 | 0.265 | 0.447 |
| Partial, PTA | L_mHG | 0.999 | 60.604 | 0.009 | 0.048 | 0.000 | 0.322 | 0.740 |
| Partial, PTA<30dB | L_mHG | 1.360 | 51.252 | 0.018 | 0.073 | 0.000 | 0.180 | 0.561 |
| Complete | R_mHG | 1.181 | 94.409 | 0.010 | 0.043 | 0.000 | 0.241 | 0.438 |
| Partial, PTA | R_mHG | 0.760 | 65.114 | 0.006 | 0.041 | 0.000 | 0.450 | 0.756 |
| Partial, PTA<30dB | R_mHG | 0.960 | 51.353 | 0.010 | 0.057 | 0.000 | 0.342 | 0.702 |
| Complete | L_MGN | -0.501 | 87.234 | 0.001 | 0.022 | 0.000 | 0.617 | 0.846 |
| Partial, PTA | L_MGN | -0.200 | 64.297 | 0.000 | 0.024 | 0.000 | 0.842 | 0.933 |
| Partial, PTA<30dB | L_MGN | -0.232 | 52.826 | 0.000 | 0.029 | 0.000 | 0.817 | 0.924 |
| Complete | R_MGN | 0.055 | 78.065 | 0.000 | 0.016 | 0.000 | 0.956 | 0.983 |
| Partial, PTA | R_MGN | -0.143 | 56.179 | 0.000 | 0.022 | 0.000 | 0.887 | 0.933 |
| Partial, PTA<30dB | R_MGN | -0.153 | 49.202 | 0.000 | 0.027 | 0.000 | 0.879 | 0.929 |
| Complete | L_IC | 1.503 | 90.189 | 0.010 | 0.043 | 0.000 | 0.136 | 0.438 |
| Partial, PTA | L_IC | 0.945 | 72.100 | 0.005 | 0.039 | 0.000 | 0.348 | 0.740 |
| Partial, PTA<30dB | L_IC | 0.998 | 64.523 | 0.006 | 0.048 | 0.000 | 0.322 | 0.701 |
| Complete | R_IC | 1.976 | 101.840 | 0.018 | 0.058 | 0.001 | 0.051 | 0.269 |
| Partial, PTA | R_IC | 1.101 | 74.387 | 0.007 | 0.045 | 0.000 | 0.275 | 0.740 |
| Partial, PTA<30dB | R_IC | 1.286 | 60.512 | 0.012 | 0.061 | 0.000 | 0.203 | 0.561 |
| ***Complete*** | ***L_dCB*** | ***3.402*** | ***92.680*** | ***0.065*** | ***0.126*** | ***0.022*** | ***0.001*** | ***0.021*** |
| Partial, PTA | L_dCB | 2.527 | 72.512 | 0.043 | 0.107 | 0.007 | 0.014 | 0.127 |
| Partial, PTA<30dB | L_dCB | 2.447 | 57.077 | 0.049 | 0.122 | 0.007 | 0.018 | 0.162 |
| ***Complete*** | ***R_dCB[t]*** | ***2.535*** | ***95.451*** | ***0.039*** | ***0.091*** | ***0.008*** | ***0.013*** | ***0.095*** |
| Partial, PTA | R_dCB | 1.375 | 68.478 | 0.016 | 0.063 | 0.000 | 0.174 | 0.740 |
| Partial, PTA<30dB | R_dCB | 1.449 | 54.719 | 0.021 | 0.079 | 0.000 | 0.153 | 0.561 |
| Complete | L_vCB | 1.162 | 92.745 | 0.006 | 0.035 | 0.000 | 0.248 | 0.438 |
| Partial, PTA | L_vCB | 1.429 | 73.629 | 0.013 | 0.057 | 0.000 | 0.157 | 0.740 |
| Partial, PTA<30dB | L_vCB | 1.191 | 57.500 | 0.011 | 0.060 | 0.000 | 0.239 | 0.589 |
| ***Complete*** | ***R_vCB[t]*** | ***2.668*** | ***89.280*** | ***0.036*** | ***0.087*** | ***0.007*** | ***0.009*** | ***0.095*** |
| ***Partial, PTA*** | ***R_vCB**** | ***3.177*** | ***63.224*** | ***0.059*** | ***0.130*** | ***0.015*** | ***0.002*** | ***0.047*** |
| ***Partial, PTA<30dB*** | ***R_vCB[t]*** | ***2.855*** | ***52.991*** | ***0.058*** | ***0.136*** | ***0.011*** | ***0.006*** | ***0.076*** |
| ***Complete*** | ***L_DN**** | ***3.362*** | ***91.384*** | ***0.040*** | ***0.092*** | ***0.008*** | ***0.001*** | ***0.021*** |
| ***Partial, PTA*** | ***L_DN**** | ***3.025*** | ***74.120*** | ***0.042*** | ***0.106*** | ***0.006*** | ***0.003*** | ***0.047*** |
| ***Partial, PTA<30dB*** | ***L_DN**** | ***3.117*** | ***65.045*** | ***0.054*** | ***0.130*** | ***0.009*** | ***0.003*** | ***0.050*** |
| Complete | R_DN | 1.255 | 106.701 | 0.007 | 0.037 | 0.000 | 0.212 | 0.438 |
| Partial, PTA | R_DN | 1.170 | 72.490 | 0.009 | 0.048 | 0.000 | 0.246 | 0.740 |
| Partial, PTA<30dB | R_DN | 1.495 | 64.014 | 0.016 | 0.069 | 0.000 | 0.140 | 0.561 |
| Complete | L_dCS | 2.153 | 98.549 | 0.025 | 0.070 | 0.002 | 0.034 | 0.208 |
| Partial, PTA | L_dCS | 1.913 | 66.795 | 0.033 | 0.091 | 0.003 | 0.060 | 0.445 |
| Partial, PTA<30dB | L_dCS | 2.076 | 53.105 | 0.044 | 0.116 | 0.005 | 0.043 | 0.316 |
| ***Complete*** | ***R_dCS[t]*** | ***2.563*** | ***85.677*** | ***0.035*** | ***0.085*** | ***0.006*** | ***0.012*** | ***0.095*** |
| ***Partial, PTA*** | ***R_dCS**** | ***3.006*** | ***61.202*** | ***0.060*** | ***0.130*** | ***0.015*** | ***0.004*** | ***0.047*** |
| ***Partial, PTA<30dB*** | ***R_dCS**** | ***3.318*** | ***46.448*** | ***0.081*** | ***0.166*** | ***0.023*** | ***0.002*** | ***0.050*** |
| Complete | L_CS | 1.839 | 109.035 | 0.025 | 0.070 | 0.002 | 0.069 | 0.282 |
| Partial, PTA | L_CS | 0.611 | 70.793 | 0.005 | 0.038 | 0.000 | 0.543 | 0.804 |
| Partial, PTA<30dB | L_CS | 0.683 | 57.410 | 0.006 | 0.048 | 0.000 | 0.497 | 0.710 |
| Complete | R_CS | 1.865 | 100.314 | 0.029 | 0.076 | 0.004 | 0.065 | 0.282 |
| Partial, PTA | R_CS | 0.465 | 63.662 | 0.003 | 0.032 | 0.000 | 0.643 | 0.810 |
| Partial, PTA<30dB | R_CS | 0.577 | 53.423 | 0.005 | 0.044 | 0.000 | 0.566 | 0.748 |
| Complete | L_aIns | -0.327 | 106.002 | 0.001 | 0.019 | 0.000 | 0.745 | 0.861 |
| Partial, PTA | L_aIns | 0.446 | 73.608 | 0.002 | 0.030 | 0.000 | 0.657 | 0.810 |

| | | | | | | | | |
|---|---|---|---|---|---|---|---|---|
| Partial, PTA<30dB | L_aIns | 0.743 | 58.064 | 0.006 | 0.048 | 0.000 | 0.460 | 0.710 |
| Complete | L_DLPFC | -1.371 | 98.567 | 0.010 | 0.043 | 0.000 | 0.174 | 0.438 |
| Partial, PTA | L_DLPFC | -0.507 | 67.252 | 0.002 | 0.030 | 0.000 | 0.614 | 0.810 |
| Partial, PTA<30dB | L_DLPFC | -0.496 | 52.633 | 0.002 | 0.036 | 0.000 | 0.622 | 0.766 |
| Complete | L_OFC | 1.217 | 89.997 | 0.008 | 0.039 | 0.000 | 0.227 | 0.438 |
| Partial, PTA | L_OFC | 0.154 | 67.890 | 0.000 | 0.022 | 0.000 | 0.878 | 0.933 |
| Partial, PTA<30dB | L_OFC | 0.512 | 54.973 | 0.002 | 0.035 | 0.000 | 0.611 | 0.766 |
| Complete | L_PHG | 0.238 | 90.342 | 0.000 | 0.018 | 0.000 | 0.812 | 0.884 |
| Partial, PTA | L_PHG | -0.085 | 61.370 | 0.000 | 0.022 | 0.000 | 0.933 | 0.933 |
| Partial, PTA<30dB | L_PHG | 0.024 | 48.972 | 0.000 | 0.026 | 0.000 | 0.981 | 0.981 |
| Complete | L_Th | 1.290 | 86.509 | 0.007 | 0.038 | 0.000 | 0.200 | 0.438 |
| Partial, PTA | L_Th | 0.853 | 70.279 | 0.004 | 0.038 | 0.000 | 0.397 | 0.740 |
| Partial, PTA<30dB | L_Th | 0.832 | 58.360 | 0.005 | 0.045 | 0.000 | 0.409 | 0.709 |
| Complete | L_VC | 0.765 | 91.241 | 0.003 | 0.027 | 0.000 | 0.446 | 0.660 |
| Partial, PTA | L_VC | 0.520 | 60.499 | 0.002 | 0.030 | 0.000 | 0.605 | 0.810 |
| Partial, PTA<30dB | L_VC | 0.468 | 55.316 | 0.002 | 0.034 | 0.000 | 0.642 | 0.766 |
| Complete | L_VS | 1.222 | 112.700 | 0.006 | 0.036 | 0.000 | 0.224 | 0.438 |
| Partial, PTA | L_VS | 0.988 | 75.118 | 0.006 | 0.042 | 0.000 | 0.326 | 0.740 |
| Partial, PTA<30dB | L_VS | 1.260 | 60.612 | 0.012 | 0.060 | 0.000 | 0.212 | 0.561 |
| Complete | preSMA | -0.832 | 101.010 | 0.004 | 0.030 | 0.000 | 0.407 | 0.628 |
| Partial, PTA | preSMA | -0.390 | 69.373 | 0.001 | 0.027 | 0.000 | 0.698 | 0.833 |
| Partial, PTA<30dB | preSMA | -0.223 | 58.269 | 0.000 | 0.028 | 0.000 | 0.824 | 0.924 |
| Complete | R_aIns | 0.863 | 101.821 | 0.004 | 0.031 | 0.000 | 0.390 | 0.628 |
| Partial, PTA | R_aIns | 0.554 | 71.390 | 0.002 | 0.031 | 0.000 | 0.581 | 0.810 |
| Partial, PTA<30dB | R_aIns | 0.827 | 60.997 | 0.006 | 0.047 | 0.000 | 0.411 | 0.709 |
| Complete | R_DLPFC | -1.639 | 98.436 | 0.013 | 0.050 | 0.000 | 0.104 | 0.386 |
| Partial, PTA | R_DLPFC | -1.068 | 66.098 | 0.009 | 0.049 | 0.000 | 0.289 | 0.740 |
| Partial, PTA<30dB | R_DLPFC | -1.397 | 50.282 | 0.018 | 0.074 | 0.000 | 0.169 | 0.561 |
| Complete | R_IFG | 0.597 | 84.024 | 0.002 | 0.023 | 0.000 | 0.552 | 0.786 |
| Partial, PTA | R_IFG | -0.829 | 58.576 | 0.005 | 0.038 | 0.000 | 0.410 | 0.740 |
| Partial, PTA<30dB | R_IFG | -0.651 | 50.850 | 0.003 | 0.038 | 0.000 | 0.518 | 0.710 |
| Complete | R_OFC | -0.260 | 100.705 | 0.000 | 0.018 | 0.000 | 0.795 | 0.884 |
| Partial, PTA | R_OFC | -0.933 | 76.275 | 0.005 | 0.039 | 0.000 | 0.354 | 0.740 |
| Partial, PTA<30dB | R_OFC | -0.809 | 65.826 | 0.004 | 0.043 | 0.000 | 0.421 | 0.709 |
| Complete | R_PHG | 0.360 | 85.342 | 0.001 | 0.019 | 0.000 | 0.720 | 0.859 |
| Partial, PTA | R_PHG | -1.076 | 71.941 | 0.007 | 0.044 | 0.000 | 0.285 | 0.740 |
| Partial, PTA<30dB | R_PHG | -0.877 | 61.828 | 0.005 | 0.045 | 0.000 | 0.384 | 0.709 |
| Complete | R_Th | 1.222 | 81.938 | 0.008 | 0.040 | 0.000 | 0.225 | 0.438 |
| Partial, PTA | R_Th | 0.630 | 60.629 | 0.002 | 0.032 | 0.000 | 0.531 | 0.804 |
| Partial, PTA<30dB | R_Th | 0.753 | 54.179 | 0.004 | 0.042 | 0.000 | 0.455 | 0.710 |
| Complete | R_VC | 0.383 | 89.010 | 0.001 | 0.020 | 0.000 | 0.702 | 0.859 |
| Partial, PTA | R_VC | -0.867 | 60.368 | 0.005 | 0.039 | 0.000 | 0.389 | 0.740 |
| Partial, PTA<30dB | R_VC | -1.750 | 60.193 | 0.020 | 0.077 | 0.000 | 0.085 | 0.525 |
| Complete | R_VS | 0.468 | 80.880 | 0.001 | 0.020 | 0.000 | 0.641 | 0.847 |
| Partial, PTA | R_VS | 0.813 | 58.283 | 0.004 | 0.036 | 0.000 | 0.420 | 0.740 |
| Partial, PTA<30dB | R_VS | 1.086 | 51.610 | 0.008 | 0.051 | 0.000 | 0.282 | 0.653 |
| Complete | rACC | -1.327 | 91.162 | 0.008 | 0.039 | 0.000 | 0.188 | 0.438 |
| Partial, PTA | rACC | -0.611 | 65.823 | 0.002 | 0.031 | 0.000 | 0.543 | 0.804 |
| Partial, PTA<30dB | rACC | -0.655 | 55.151 | 0.003 | 0.039 | 0.000 | 0.515 | 0.710 |

**Nodes pFDR < 0.05 and nodes ${}^{t}$pFDR < 0.10 are marked in bold italic font.*

**Table S6. Tinnitus Impact and Auditory Network Connectivity**

| FC Definition | roi | t | df | r2 | r2_uCI | r2_lCI | p | $p_{FDR}$ |
|---|---|---|---|---|---|---|---|---|
| HG/HS Seed | netStrength | 0.679 | 35.120 | 0.010 | 0.087 | 0.000 | 0.501 | NA |
| HG/HS Seed | L_HG | -0.664 | 36.355 | 0.013 | 0.092 | 0.000 | 0.511 | 0.787 |
| HG/HS Seed | R_HG | 1.532 | 34.749 | 0.055 | 0.169 | 0.002 | 0.135 | 0.478 |
| HG/HS Seed | L_lHG | -0.827 | 34.410 | 0.015 | 0.097 | 0.000 | 0.414 | 0.730 |
| HG/HS Seed | R_lHG | 1.834 | 37.091 | 0.078 | 0.202 | 0.008 | 0.075 | 0.460 |
| HG/HS Seed | L_mHG | 0.169 | 36.095 | 0.001 | 0.054 | 0.000 | 0.867 | 0.915 |
| HG/HS Seed | R_mHG | 1.446 | 34.989 | 0.056 | 0.172 | 0.002 | 0.157 | 0.478 |
| HG/HS Seed | L_MGN | 0.666 | 33.127 | 0.005 | 0.071 | 0.000 | 0.510 | 0.787 |
| HG/HS Seed | R_MGN | -0.326 | 37.005 | 0.002 | 0.059 | 0.000 | 0.746 | 0.892 |
| HG/HS Seed | L_IC | -1.089 | 30.186 | 0.019 | 0.107 | 0.000 | 0.285 | 0.654 |
| HG/HS Seed | R_IC | -0.340 | 32.257 | 0.002 | 0.061 | 0.000 | 0.736 | 0.892 |
| HG/HS Seed | L_dCB* | -3.843 | 35.452 | 0.191 | 0.335 | 0.074 | 0.000 | 0.018 |
| HG/HS Seed | R_dCB[t] | -2.859 | 34.759 | 0.124 | 0.261 | 0.030 | 0.007 | 0.088 |
| HG/HS Seed | L_vCB | -2.343 | 37.037 | 0.108 | 0.243 | 0.021 | 0.025 | 0.228 |
| HG/HS Seed | R_vCB | -1.409 | 34.304 | 0.034 | 0.137 | 0.000 | 0.168 | 0.478 |
| HG/HS Seed | L_DN | -0.981 | 91.000 | 0.010 | 0.087 | 0.000 | 0.329 | 0.654 |
| HG/HS Seed | R_DN | 1.528 | 20.287 | 0.026 | 0.121 | 0.000 | 0.142 | 0.478 |
| HG/HS Seed | L_dCS | 0.878 | 32.396 | 0.011 | 0.088 | 0.000 | 0.386 | 0.715 |
| HG/HS Seed | R_dCS | 0.990 | 35.159 | 0.015 | 0.099 | 0.000 | 0.329 | 0.654 |
| HG/HS Seed | L_CS | 0.324 | 32.536 | 0.002 | 0.059 | 0.000 | 0.748 | 0.892 |
| HG/HS Seed | R_CS | 1.605 | 22.071 | 0.039 | 0.145 | 0.000 | 0.123 | 0.478 |
| HG/HS Seed | L_Th | -0.103 | 36.517 | 0.000 | 0.052 | 0.000 | 0.918 | 0.918 |
| HG/HS Seed | R_Th | -0.481 | 33.031 | 0.005 | 0.070 | 0.000 | 0.634 | 0.892 |
| HG/HS Seed | L_aIns | 0.341 | 31.986 | 0.002 | 0.062 | 0.000 | 0.735 | 0.892 |
| HG/HS Seed | R_aIns | 0.976 | 32.437 | 0.019 | 0.107 | 0.000 | 0.336 | 0.654 |
| HG/HS Seed | L_DLPFC | 0.776 | 33.281 | 0.012 | 0.091 | 0.000 | 0.443 | 0.746 |
| HG/HS Seed | R_DLPFC | 0.441 | 35.404 | 0.004 | 0.069 | 0.000 | 0.662 | 0.892 |
| HG/HS Seed | L_OFC | -0.406 | 35.942 | 0.004 | 0.068 | 0.000 | 0.687 | 0.892 |
| HG/HS Seed | R_OFC[t] | 3.018 | 35.353 | 0.148 | 0.291 | 0.044 | 0.005 | 0.087 |
| HG/HS Seed | R_IFG | 0.227 | 24.937 | 0.001 | 0.056 | 0.000 | 0.822 | 0.915 |
| HG/HS Seed | L_PHG | 1.451 | 91.000 | 0.022 | 0.114 | 0.000 | 0.150 | 0.478 |
| HG/HS Seed | R_PHG | 1.545 | 31.102 | 0.038 | 0.144 | 0.000 | 0.132 | 0.478 |
| HG/HS Seed | L_VC | -0.293 | 29.968 | 0.001 | 0.057 | 0.000 | 0.771 | 0.892 |
| HG/HS Seed | R_VC | -0.174 | 29.202 | 0.000 | 0.054 | 0.000 | 0.863 | 0.915 |
| HG/HS Seed | L_VS | 1.910 | 33.046 | 0.060 | 0.178 | 0.003 | 0.065 | 0.460 |
| HG/HS Seed | R_VS | 1.345 | 31.892 | 0.027 | 0.123 | 0.000 | 0.188 | 0.497 |
| HG/HS Seed | preSMA | 1.070 | 34.613 | 0.019 | 0.107 | 0.000 | 0.292 | 0.654 |
| HG/HS Seed | rACC | 0.139 | 33.998 | 0.000 | 0.053 | 0.000 | 0.890 | 0.915 |
| Yeo17 DR | netStrength | 0.028 | 35.978 | 0.000 | 0.050 | 0.000 | 0.978 | NA |
| Yeo17 DR | L_HG | -0.254 | 32.924 | 0.002 | 0.056 | 0.000 | 0.801 | 0.945 |
| Yeo17 DR | R_HG | 0.731 | 35.652 | 0.015 | 0.095 | 0.000 | 0.470 | 0.869 |
| Yeo17 DR | L_lHG | -1.551 | 30.821 | 0.044 | 0.149 | 0.001 | 0.131 | 0.779 |
| Yeo17 DR | R_lHG | 0.321 | 35.081 | 0.003 | 0.061 | 0.000 | 0.750 | 0.945 |
| Yeo17 DR | L_mHG | 0.200 | 31.795 | 0.001 | 0.054 | 0.000 | 0.843 | 0.945 |
| Yeo17 DR | R_mHG | 1.397 | 36.392 | 0.051 | 0.160 | 0.002 | 0.171 | 0.790 |
| Yeo17 DR | L_MGN | -0.297 | 31.632 | 0.002 | 0.057 | 0.000 | 0.768 | 0.945 |
| Yeo17 DR | R_MGN | 0.334 | 29.929 | 0.002 | 0.058 | 0.000 | 0.741 | 0.945 |
| Yeo17 DR | L_IC | -1.800 | 33.462 | 0.043 | 0.148 | 0.001 | 0.081 | 0.779 |
| Yeo17 DR | R_IC | -0.024 | 35.259 | 0.000 | 0.049 | 0.000 | 0.981 | 0.981 |
| Yeo17 DR | L_dCB | -1.240 | 27.589 | 0.024 | 0.116 | 0.000 | 0.225 | 0.869 |
| Yeo17 DR | R_dCB | -0.370 | 28.834 | 0.003 | 0.063 | 0.000 | 0.714 | 0.945 |
| Yeo17 DR | L_vCB | -1.593 | 31.702 | 0.027 | 0.121 | 0.000 | 0.121 | 0.779 |
| Yeo17 DR | R_vCB | -0.683 | 22.689 | 0.005 | 0.069 | 0.000 | 0.501 | 0.883 |
| Yeo17 DR | L_DN | 0.791 | 22.985 | 0.007 | 0.076 | 0.000 | 0.437 | 0.869 |
| Yeo17 DR | R_DN | 2.474 | 33.166 | 0.111 | 0.242 | 0.024 | 0.019 | 0.691 |
| Yeo17 DR | L_dCS | 0.810 | 35.651 | 0.013 | 0.091 | 0.000 | 0.424 | 0.869 |
| Yeo17 DR | R_dCS | 1.480 | 36.614 | 0.040 | 0.143 | 0.001 | 0.147 | 0.779 |
| Yeo17 DR | L_CS | -0.968 | 38.111 | 0.032 | 0.131 | 0.000 | 0.339 | 0.869 |
| Yeo17 DR | R_CS | -0.382 | 31.814 | 0.004 | 0.065 | 0.000 | 0.705 | 0.945 |
| Yeo17 DR | L_Th | 0.321 | 95.000 | 0.001 | 0.055 | 0.000 | 0.749 | 0.945 |
| Yeo17 DR | R_Th | 0.251 | 29.096 | 0.001 | 0.055 | 0.000 | 0.804 | 0.945 |
| Yeo17 DR | L_aIns | 0.746 | 37.386 | 0.013 | 0.090 | 0.000 | 0.461 | 0.869 |

| | | | | | | | | |
|---|---|---|---|---|---|---|---|---|
| Yeo17 DR | R_aIns | 0.901 | 34.603 | 0.013 | 0.093 | 0.000 | 0.374 | 0.869 |
| Yeo17 DR | L_DLPFC | 0.045 | 33.194 | 0.000 | 0.050 | 0.000 | 0.964 | 0.981 |
| Yeo17 DR | R_DLPFC | -0.749 | 32.999 | 0.007 | 0.077 | 0.000 | 0.459 | 0.869 |
| Yeo17 DR | L_OFC | 0.052 | 28.947 | 0.000 | 0.050 | 0.000 | 0.959 | 0.981 |
| Yeo17 DR | R_OFC | 1.056 | 31.497 | 0.013 | 0.091 | 0.000 | 0.299 | 0.869 |
| Yeo17 DR | R_IFG | 0.565 | 34.350 | 0.008 | 0.078 | 0.000 | 0.576 | 0.945 |
| Yeo17 DR | L_PHG | 0.223 | 30.994 | 0.001 | 0.054 | 0.000 | 0.825 | 0.945 |
| Yeo17 DR | R_PHG | -0.425 | 30.042 | 0.002 | 0.057 | 0.000 | 0.674 | 0.945 |
| Yeo17 DR | L_VC | -0.804 | 32.859 | 0.015 | 0.096 | 0.000 | 0.427 | 0.869 |
| Yeo17 DR | R_VC | -1.682 | 28.284 | 0.046 | 0.153 | 0.001 | 0.104 | 0.779 |
| Yeo17 DR | L_VS | 1.091 | 36.464 | 0.017 | 0.102 | 0.000 | 0.282 | 0.869 |
| Yeo17 DR | R_VS | -0.072 | 29.401 | 0.000 | 0.050 | 0.000 | 0.943 | 0.981 |
| Yeo17 DR | preSMA | 1.060 | 36.246 | 0.023 | 0.113 | 0.000 | 0.296 | 0.869 |
| Yeo17 DR | rACC | -1.619 | 24.979 | 0.034 | 0.135 | 0.000 | 0.118 | 0.779 |

**Nodes pFDR < 0.05 and nodes [t]pFDR < 0.10 are marked in bold italic font.*

**Table S7. LTA-CtDCS HG/HS Seed Connectivity**

| | Main Effect | | | | Sham vs. Active | | Rest vs. Active | | Rest vs. Inactive | |
|---|---|---|---|---|---|---|---|---|---|---|
| roi | F | DF | p | $p_{FDR}$ | Cohen’s d (CI) | p | Cohen’s d (CI) | p | Cohen’s d (CI) | p |
| ***netStrength**** | ***15.14*** | ***2,61*** | ***0*** | ***0*** | ***-0.69 (-1.21, -0.17)*** | ***0.009*** | ***-1.47 (-2.05, -0.89)*** | ***0*** | ***-0.78(-1.37, -0.19)*** | ***0.008*** |
| L_HG | 7.6 | 2,62 | 0.001 | 0.01 | -0.15(-0.66, 0.36) | 0.552 | -1.01(-1.57, -0.45) | 0 | -0.86(-1.45, -0.27) | 0.004 |
| R_HG | 10.55 | 2,61 | 0 | 0.001 | -0.38(-0.9, 0.13) | 0.132 | -1.23(-1.8, -0.66) | 0 | -0.85(-1.44, -0.25) | 0.004 |
| L_lHG | 4.45 | 2,63 | 0.016 | 0.064 | -0.18(-0.68, 0.32) | 0.472 | -0.79(-1.34, -0.24) | 0.005 | -0.61(-1.19, -0.03) | 0.037 |
| R_lHG | 3.49 | 2,60 | 0.037 | 0.108 | -0.01(-0.53, 0.51) | 0.976 | -0.65(-1.21, -0.1) | 0.018 | -0.65(-1.23, -0.06) | 0.028 |
| L_mHG | 4.28 | 2,62 | 0.018 | 0.064 | 0.07(-0.43, 0.57) | 0.770 | -0.69(-1.23, -0.14) | 0.013 | -0.76(-1.34, -0.18) | 0.01 |
| R_mHG | 7.44 | 2,60 | 0.001 | 0.01 | -0.04(-0.55, 0.47) | 0.867 | -0.98(-1.55, -0.41) | 0.001 | -0.94(-1.53, -0.34) | 0.002 |
| L_MGN | 2.84 | 2,61 | 0.066 | 0.168 | 0.02(-0.47, 0.52) | 0.924 | -0.58(-1.13, -0.03) | 0.035 | -0.61(-1.19, -0.02) | 0.039 |
| R_MGN | 1.33 | 2,61 | 0.271 | 0.448 | -0.07(-0.57, 0.43) | 0.782 | -0.43(-0.96, 0.11) | 0.118 | -0.36(-0.93, 0.22) | 0.217 |
| L_IC | 0.94 | 2,62 | 0.397 | 0.584 | -0.3(-0.81, 0.2) | 0.230 | -0.29(-0.84, 0.25) | 0.288 | 0.01(-0.56, 0.58) | 0.965 |
| R_IC | 1.57 | 2,61 | 0.216 | 0.411 | 0.16(-0.34, 0.67) | 0.526 | -0.34(-0.89, 0.21) | 0.217 | -0.5(-1.08, 0.07) | 0.085 |
| L_dCB | 0.45 | 2,62 | 0.639 | 0.809 | -0.21(-0.72, 0.29) | 0.397 | -0.2(-0.74, 0.35) | 0.471 | 0.02(-0.56, 0.59) | 0.956 |
| R_dCB | 1.2 | 2,61 | 0.309 | 0.49 | -0.3(-0.8, 0.2) | 0.230 | -0.36(-0.9, 0.17) | 0.178 | -0.06(-0.63, 0.51) | 0.827 |
| L_vCB | 1.47 | 2,62 | 0.238 | 0.43 | -0.12(-0.61, 0.38) | 0.639 | -0.46(-1, 0.08) | 0.094 | -0.34(-0.92, 0.23) | 0.238 |
| R_vCB | 0.34 | 2,89 | 0.709 | 0.817 | -0.09(-0.59, 0.41) | 0.713 | -0.22(-0.76, 0.31) | 0.411 | -0.13(-0.71, 0.45) | 0.658 |
| L_DN | 2.07 | 2,63 | 0.135 | 0.301 | -0.41(-0.91, 0.09) | 0.101 | -0.46(-1, 0.07) | 0.088 | -0.05(-0.62, 0.52) | 0.86 |
| R_DN | 0.18 | 2,61 | 0.84 | 0.886 | -0.07(-0.59, 0.44) | 0.771 | -0.16(-0.69, 0.38) | 0.558 | -0.08(-0.65, 0.49) | 0.772 |
| L_dCS | 6.5 | 2,63 | 0.003 | 0.015 | 0.05(-0.45, 0.54) | 0.847 | -0.87(-1.41, -0.32) | 0.002 | -0.91(-1.5, -0.33) | 0.002 |
| R_dCS | 5.36 | 2,60 | 0.007 | 0.034 | -0.02(-0.53, 0.48) | 0.924 | -0.82(-1.38, -0.27) | 0.003 | -0.8(-1.38, -0.21) | 0.007 |
| L_CS | 11.04 | 2,59 | 0 | 0.001 | -0.23(-0.73, 0.28) | 0.378 | -1.24(-1.82, -0.67) | 0 | -1.02(-1.62, -0.42) | 0.001 |
| R_CS | 6.56 | 2,59 | 0.003 | 0.015 | -0.15(-0.66, 0.36) | 0.548 | -0.97(-1.54, -0.39) | 0.001 | -0.81(-1.4, -0.22) | 0.007 |
| ***L_Th[t]*** | ***4.26*** | ***2,62*** | ***0.018*** | ***0.064*** | ***-0.7(-1.22, -0.19)*** | ***0.007*** | ***-0.52(-1.06, 0.02)*** | ***0.056*** | ***0.18(-0.39, 0.75)*** | ***0.534*** |
| R_Th | 0.73 | 2,60 | 0.488 | 0.662 | -0.17(-0.68, 0.33) | 0.493 | -0.32(-0.87, 0.22) | 0.241 | -0.15(-0.72, 0.42) | 0.606 |
| L_aIns | 2.59 | 2,59 | 0.083 | 0.198 | -0.33(-0.83, 0.18) | 0.197 | -0.61(-1.17, -0.06) | 0.029 | -0.28(-0.86, 0.29) | 0.326 |
| R_aIns | 0.4 | 2,62 | 0.671 | 0.817 | -0.06(-0.56, 0.45) | 0.822 | -0.24(-0.78, 0.3) | 0.379 | -0.18(-0.76, 0.39) | 0.528 |
| L_DLPFC | 1.6 | 2,63 | 0.211 | 0.411 | -0.44(-0.95, 0.06) | 0.079 | -0.17(-0.7, 0.37) | 0.532 | 0.28(-0.29, 0.85) | 0.336 |
| R_DLPFC | 0.01 | 2,62 | 0.989 | 0.99 | -0.01(-0.51, 0.49) | 0.977 | 0.03(-0.51, 0.58) | 0.902 | 0.04(-0.53, 0.61) | 0.886 |
| L_OFC | 0.35 | 2,63 | 0.707 | 0.817 | -0.21(-0.71, 0.29) | 0.407 | -0.09(-0.62, 0.45) | 0.751 | 0.12(-0.45, 0.69) | 0.669 |
| R_OFC | 0.78 | 2,61 | 0.464 | 0.653 | -0.12(-0.63, 0.38) | 0.633 | 0.23(-0.32, 0.78) | 0.396 | 0.35(-0.22, 0.93) | 0.22 |
| R_IFG | 0.93 | 2,62 | 0.399 | 0.584 | -0.1(-0.6, 0.4) | 0.698 | -0.36(-0.9, 0.17) | 0.179 | -0.27(-0.85, 0.31) | 0.363 |
| L_PHG | 0.21 | 2,63 | 0.807 | 0.885 | -0.01(-0.52, 0.49) | 0.953 | 0.16(-0.38, 0.69) | 0.563 | 0.17(-0.41, 0.75) | 0.559 |
| R_PHG | 1.35 | 2,62 | 0.267 | 0.448 | -0.23(-0.73, 0.28) | 0.373 | 0.24(-0.29, 0.78) | 0.368 | 0.47(-0.11, 1.04) | 0.106 |
| L_VC | 0.01 | 2,58 | 0.99 | 0.99 | 0.03(-0.48, 0.54) | 0.895 | 0(-0.55, 0.56) | 0.988 | -0.03(-0.6, 0.54) | 0.918 |
| R_VC | 0.67 | 2,55 | 0.516 | 0.677 | 0.28(-0.23, 0.8) | 0.279 | 0.03(-0.52, 0.57) | 0.918 | -0.25(-0.83, 0.32) | 0.383 |
| L_VS | 3.73 | 2,59 | 0.03 | 0.094 | -0.59(-1.11, -0.08) | 0.022 | -0.61(-1.16, -0.06) | 0.03 | -0.01(-0.58, 0.56) | 0.965 |
| R_VS | 1.68 | 2,62 | 0.195 | 0.411 | -0.44(-0.94, 0.07) | 0.087 | -0.33(-0.88, 0.21) | 0.223 | 0.1(-0.47, 0.67) | 0.723 |
| preSMA | 3.15 | 2,62 | 0.05 | 0.135 | -0.52(-1.03, -0.01) | 0.043 | -0.57(-1.11, -0.03) | 0.037 | -0.05(-0.62, 0.52) | 0.856 |
| rACC | 0.21 | 2,61 | 0.815 | 0.885 | 0.1(-0.4, 0.6) | 0.689 | -0.08(-0.62, 0.46) | 0.764 | -0.18(-0.75, 0.39) | 0.529 |

**Main effect pfdr < 0.05 AND sham vs. active p < 0.05; tMain effect pfdr<010 AND sham vs. active p < 0.05.*
*P values listed as “0” are p < 0.001*
*For pairwise comparisons, negative Cohen's d indicates that first condition listed is less than second condition (e.g., NetStrength is less for sham than active and Cohen's d is negative).*

**Table S8. STC-CtDCS HG/HS Seed Connectivity**

| | Main Effect | | | | Sham vs. Active | | Rest vs. Active | | Rest vs. Inactive | |
|---|---|---|---|---|---|---|---|---|---|---|
| roi | F | DF | p | pFDR | Cohen's d (CI) | p | Cohen's d (CI) | p | Cohen's d (CI) | p |
| netStrength | 0.62 | 2,16 | 0.551 | 0.91 | -0.57(-2.71, 1.57) | 0.55 | -0.06(-2.01, 1.88) | 0.94 | 0.5(-0.57, 1.57) | 0.29 |
| L_HG | 2.46 | 2,13 | 0.123 | 0.585 | -0.7(-2.95, 1.56) | 0.483 | 0.36(-1.76, 2.48) | 0.698 | 1.06(-0.12, 2.23) | 0.045 |
| R_HG | 2.36 | 2,19 | 0.122 | 0.585 | -1.5(-3.46, 0.47) | 0.094 | -0.67(-2.44, 1.09) | 0.398 | 0.82(-0.24, 1.89) | 0.09 |
| L_lHG | 1.03 | 2,17 | 0.38 | 0.759 | -1.27(-3.46, 0.92) | 0.189 | -1.09(-3.08, 0.9) | 0.212 | 0.18(-0.88, 1.24) | 0.702 |
| R_lHG | 1.03 | 2,18 | 0.377 | 0.759 | -1.18(-3.21, 0.86) | 0.197 | -0.79(-2.64, 1.07) | 0.341 | 0.39(-0.65, 1.44) | 0.402 |
| L_mHG | 1.29 | 2,19 | 0.3 | 0.67 | -0.73(-2.45, 1) | 0.366 | -0.01(-1.6, 1.57) | 0.985 | 0.71(-0.31, 1.74) | 0.136 |
| R_mHG | 2.47 | 2,18 | 0.112 | 0.585 | -0.49(-2.43, 1.45) | 0.575 | 0.51(-1.28, 2.3) | 0.528 | 1(-0.09, 2.09) | 0.044 |
| L_MGN | 1.42 | 2,18 | 0.267 | 0.649 | 1.48(-0.67, 3.63) | 0.122 | 1.16(-0.79, 3.11) | 0.178 | -0.32(-1.37, 0.74) | 0.501 |
| R_MGN | 0.14 | 2,17 | 0.872 | 0.964 | -0.33(-2.4, 1.73) | 0.719 | -0.11(-2, 1.78) | 0.896 | 0.22(-0.83, 1.27) | 0.634 |
| L_IC | 1.85 | 2,18 | 0.186 | 0.649 | 0.25(-1.69, 2.19) | 0.778 | 0.98(-0.86, 2.81) | 0.234 | 0.73(-0.34, 1.79) | 0.131 |
| R_IC | 1.71 | 2,18 | 0.208 | 0.649 | -1.05(-3.13, 1.02) | 0.257 | -0.24(-2.12, 1.63) | 0.772 | 0.81(-0.27, 1.9) | 0.096 |
| L_dCB | 1.49 | 2,15 | 0.256 | 0.649 | 0.95(-1.25, 3.15) | 0.332 | 1.4(-0.76, 3.56) | 0.143 | 0.45(-0.68, 1.59) | 0.369 |
| R_dCB | 2.75 | 2,17 | 0.092 | 0.585 | 0.33(-1.79, 2.45) | 0.727 | 1.22(-0.77, 3.2) | 0.165 | 0.89(-0.22, 1.99) | 0.072 |
| L_vCB | 0.2 | 2,17 | 0.818 | 0.964 | -0.32(-2.21, 1.58) | 0.715 | -0.45(-2.2, 1.31) | 0.571 | -0.13(-1.16, 0.89) | 0.776 |
| R_vCB | 1.86 | 2,16 | 0.189 | 0.649 | 0.11(-2.01, 2.24) | 0.903 | 0.89(-1.08, 2.85) | 0.306 | 0.77(-0.32, 1.86) | 0.112 |
| L_DN | 1.55 | 2,18 | 0.238 | 0.649 | -1.49(-3.52, 0.53) | 0.111 | -1.07(-2.93, 0.79) | 0.205 | 0.42(-0.63, 1.48) | 0.368 |
| R_DN | 0.06 | 2,17 | 0.943 | 0.964 | -0.29(-2.41, 1.83) | 0.757 | -0.19(-2.12, 1.75) | 0.828 | 0.11(-0.95, 1.16) | 0.821 |
| L_dCS | 0.24 | 2,15 | 0.788 | 0.964 | 0.01(-2.2, 2.22) | 0.991 | -0.29(-2.24, 1.67) | 0.739 | -0.3(-1.45, 0.86) | 0.56 |
| R_dCS | 0.17 | 2,17 | 0.843 | 0.964 | -0.51(-2.65, 1.64) | 0.593 | -0.33(-2.28, 1.62) | 0.702 | 0.18(-0.88, 1.24) | 0.704 |
| L_CS | 0.04 | 2,14 | 0.96 | 0.964 | 0.13(-1.89, 2.15) | 0.885 | 0(-1.86, 1.86) | 0.999 | -0.13(-1.17, 0.91) | 0.78 |
| R_CS | 0.2 | 2,17 | 0.818 | 0.964 | 0.41(-1.73, 2.54) | 0.666 | 0.5(-1.45, 2.45) | 0.558 | 0.1(-0.96, 1.15) | 0.837 |
| L_Th | 0.35 | 2,10 | 0.713 | 0.964 | 0.66(-1.35, 2.66) | 0.465 | 0.4(-1.51, 2.31) | 0.635 | -0.26(-1.35, 0.83) | 0.593 |
| R_Th | 0.41 | 2,13 | 0.671 | 0.945 | 0.44(-1.77, 2.66) | 0.649 | 0.02(-2.07, 2.12) | 0.98 | -0.42(-1.54, 0.7) | 0.391 |
| L_aIns | 0.51 | 2,18 | 0.609 | 0.945 | -0.74(-2.81, 1.32) | 0.422 | -0.79(-2.69, 1.11) | 0.346 | -0.05(-1.1, 0.99) | 0.912 |
| R_aIns | 0.89 | 2,18 | 0.428 | 0.765 | -0.66(-2.66, 1.34) | 0.462 | -0.95(-2.82, 0.91) | 0.25 | -0.29(-1.34, 0.75) | 0.533 |
| L_DLPFC | 2.66 | 2,22 | 0.093 | 0.585 | -1.67(-3.42, 0.08) | 0.04 | -1.1(-2.68, 0.48) | 0.132 | 0.57(-0.44, 1.58) | 0.228 |
| R_DLPFC | 3.67 | 2,15 | 0.05 | 0.585 | 2.04(-0.21, 4.29) | 0.058 | 2.28(0.21, 4.35) | 0.018 | 0.24(-0.86, 1.34) | 0.637 |
| L_OFC | 2.78 | 2,18 | 0.089 | 0.585 | -1.88(-3.88, 0.11) | 0.038 | -1.33(-3.13, 0.48) | 0.102 | 0.56(-0.49, 1.6) | 0.241 |
| R_OFC | 2.02 | 2,16 | 0.166 | 0.649 | -1.73(-3.85, 0.39) | 0.093 | -1.62(-3.48, 0.25) | 0.065 | 0.12(-1.01, 1.24) | 0.817 |
| R_IFG | 0.43 | 2,16 | 0.656 | 0.945 | 0.01(-1.91, 1.93) | 0.994 | -0.4(-2.23, 1.43) | 0.631 | -0.4(-1.47, 0.66) | 0.404 |
| L_PHG | 0.88 | 2,18 | 0.43 | 0.765 | -0.7(-2.72, 1.32) | 0.439 | -0.97(-2.84, 0.91) | 0.245 | -0.27(-1.31, 0.78) | 0.567 |
| R_PHG | 0.43 | 2,17 | 0.658 | 0.945 | -0.61(-2.68, 1.47) | 0.512 | -0.73(-2.62, 1.17) | 0.392 | -0.12(-1.17, 0.93) | 0.803 |
| L_VC | 0.85 | 2,18 | 0.443 | 0.765 | 0.86(-1.16, 2.89) | 0.343 | 1(-0.87, 2.87) | 0.23 | 0.14(-0.9, 1.18) | 0.766 |
| R_VC | 0.07 | 2,18 | 0.934 | 0.964 | 0.32(-1.75, 2.38) | 0.731 | 0.27(-1.62, 2.16) | 0.748 | -0.05(-1.1, 1) | 0.919 |
| L_VS | 1.4 | 2,18 | 0.273 | 0.649 | -1.33(-3.29, 0.63) | 0.134 | -1.13(-2.93, 0.67) | 0.162 | 0.2(-0.83, 1.23) | 0.667 |
| R_VS | 2.51 | 2,17 | 0.112 | 0.585 | -1.65(-3.46, 0.16) | 0.048 | -1.34(-3.01, 0.32) | 0.078 | 0.31(-0.7, 1.31) | 0.511 |
| preSMA | 0.04 | 2,17 | 0.964 | 0.964 | 0.15(-1.97, 2.28) | 0.872 | 0.21(-1.73, 2.15) | 0.808 | 0.06(-1, 1.11) | 0.906 |
| rACC | 0.21 | 2,15 | 0.812 | 0.964 | 0.15(-1.88, 2.19) | 0.868 | 0.38(-1.5, 2.25) | 0.651 | 0.22(-0.82, 1.27) | 0.631 |

**Main effect pfdr < 0.05 AND sham vs. active p < 0.05; tMain effect pfdr<010 AND sham vs. active p < 0.05.*
*P values listed as "0" are p < 0.001.*
*CI, 95% Confidence Interval.*
*For pairwise comparisons, negative Cohen's d indicates that first condition listed is less than second condition (e.g., NetStrength is less for sham than active if Cohen's d is negative).*

**Table S9. DLPFC-CtDCS HG/HS Seed Connectivity**

| | Main Effect | | | | Sham vs. Active | | Rest vs. Active | | Rest vs. Inactive | |
|---|---|---|---|---|---|---|---|---|---|---|
| roi | F | DF | p | pFDR | Cohen's d (CI) | p | Cohen's d (CI) | p | Cohen's d (CI) | p |
| netStrength | 3.15 | 2,66 | 0.05 | 0.175 | -0.65(-1.2, -0.1) | 0.018 | -0.22(-0.75, 0.3) | 0.386 | 0.42(-0.07, 0.92) | 0.086 |
| L_HG | 4.28 | 2,65 | 0.018 | 0.161 | -0.75(-1.32, -0.18) | 0.008 | -0.2(-0.73, 0.33) | 0.448 | 0.55(0.05, 1.06) | 0.028 |
| R_HG | 3.97 | 2,66 | 0.024 | 0.161 | -0.67(-1.23, -0.11) | 0.016 | -0.08(-0.6, 0.45) | 0.76 | 0.59(0.09, 1.1) | 0.018 |
| ***L_lHG***[t] | ***6.74*** | ***2,68*** | ***0.002*** | ***0.082*** | ***-0.98(-1.54, -0.42)*** | ***0*** | ***-0.59(-1.13, -0.06)*** | ***0.025*** | ***0.38(-0.1, 0.87)*** | ***0.11*** |
| R_lHG | 4.27 | 2,65 | 0.018 | 0.161 | -0.75(-1.31, -0.18) | 0.008 | -0.63(-1.17, -0.1) | 0.018 | 0.12(-0.38, 0.61) | 0.637 |
| L_mHG | 0.68 | 2,63 | 0.51 | 0.792 | -0.32(-0.88, 0.24) | 0.251 | -0.21(-0.74, 0.33) | 0.434 | 0.11(-0.39, 0.61) | 0.65 |
| R_mHG | 0.39 | 2,64 | 0.677 | 0.857 | 0.13(-0.43, 0.69) | 0.638 | -0.09(-0.61, 0.44) | 0.739 | -0.22(-0.72, 0.28) | 0.379 |
| L_MGN | 1.94 | 2,68 | 0.151 | 0.358 | -0.53(-1.08, 0.03) | 0.057 | -0.22(-0.75, 0.31) | 0.396 | 0.3(-0.19, 0.8) | 0.217 |
| R_MGN | 1.16 | 2,68 | 0.319 | 0.674 | -0.31(-0.84, 0.23) | 0.254 | 0.03(-0.49, 0.55) | 0.894 | 0.34(-0.14, 0.82) | 0.157 |
| L_IC | 4.01 | 2,68 | 0.023 | 0.161 | 0.03(-0.52, 0.58) | 0.907 | 0.61(0.09, 1.13) | 0.021 | 0.58(0.09, 1.07) | 0.02 |
| R_IC | 3.39 | 2,67 | 0.039 | 0.175 | -0.15(-0.7, 0.4) | 0.588 | 0.45(-0.08, 0.98) | 0.087 | 0.6(0.1, 1.09) | 0.016 |
| L_dCB | 0.18 | 2,66 | 0.838 | 0.965 | 0.06(-0.48, 0.61) | 0.812 | 0.15(-0.37, 0.67) | 0.561 | 0.09(-0.41, 0.58) | 0.727 |
| R_dCB | 0.64 | 2,66 | 0.528 | 0.792 | 0.31(-0.25, 0.86) | 0.266 | 0.13(-0.4, 0.66) | 0.619 | -0.17(-0.68, 0.33) | 0.488 |
| L_vCB | 0.92 | 2,67 | 0.404 | 0.733 | -0.31(-0.85, 0.23) | 0.256 | -0.33(-0.86, 0.2) | 0.212 | -0.02(-0.51, 0.46) | 0.924 |
| R_vCB | 0.12 | 2,67 | 0.885 | 0.989 | 0.07(-0.47, 0.6) | 0.801 | -0.05(-0.57, 0.47) | 0.85 | -0.12(-0.59, 0.36) | 0.622 |
| L_DN | 3.01 | 2,68 | 0.056 | 0.178 | 0.47(-0.08, 1.02) | 0.087 | -0.1(-0.62, 0.43) | 0.716 | -0.57(-1.06, -0.07) | 0.022 |
| R_DN | 0.62 | 2,68 | 0.542 | 0.792 | 0.3(-0.24, 0.83) | 0.271 | 0.18(-0.34, 0.71) | 0.484 | -0.11(-0.6, 0.37) | 0.644 |
| L_dCS | 1.1 | 2,68 | 0.338 | 0.676 | -0.39(-0.93, 0.16) | 0.159 | -0.3(-0.82, 0.22) | 0.246 | 0.08(-0.4, 0.57) | 0.728 |
| R_dCS | 3.86 | 2,68 | 0.026 | 0.161 | -0.74(-1.29, -0.19) | 0.007 | -0.4(-0.92, 0.13) | 0.128 | 0.34(-0.15, 0.84) | 0.16 |
| L_CS | 2.5 | 2,70 | 0.089 | 0.242 | -0.6(-1.15, -0.05) | 0.029 | -0.31(-0.83, 0.21) | 0.232 | 0.29(-0.19, 0.77) | 0.224 |
| R_CS | 3.11 | 2,70 | 0.051 | 0.175 | -0.6(-1.14, -0.06) | 0.026 | -0.56(-1.08, -0.04) | 0.033 | 0.04(-0.43, 0.52) | 0.858 |
| L_Th | 0 | 2,67 | 0.998 | 0.998 | 0.01(-0.54, 0.57) | 0.957 | 0(-0.52, 0.53) | 0.99 | -0.01(-0.51, 0.49) | 0.963 |
| R_Th | 0.26 | 2,67 | 0.769 | 0.914 | 0.02(-0.53, 0.57) | 0.938 | 0.16(-0.36, 0.69) | 0.529 | 0.14(-0.35, 0.63) | 0.561 |
| L_aIns | 3.7 | 2,69 | 0.03 | 0.161 | -0.61(-1.15, -0.07) | 0.024 | -0.65(-1.18, -0.12) | 0.014 | -0.04(-0.51, 0.44) | 0.876 |
| R_aIns | 3.22 | 2,70 | 0.046 | 0.175 | -0.56(-1.1, -0.01) | 0.039 | -0.61(-1.14, -0.09) | 0.019 | -0.06(-0.53, 0.42) | 0.809 |
| L_DLPFC | 0.47 | 2,69 | 0.625 | 0.849 | 0.23(-0.31, 0.77) | 0.387 | 0.04(-0.48, 0.56) | 0.872 | -0.19(-0.67, 0.29) | 0.427 |
| R_DLPFC | 0.04 | 2,69 | 0.959 | 0.998 | 0.05(-0.48, 0.59) | 0.844 | -0.01(-0.53, 0.5) | 0.956 | -0.07(-0.55, 0.41) | 0.78 |
| L_OFC | 0.67 | 2,67 | 0.516 | 0.792 | 0.29(-0.25, 0.83) | 0.282 | 0.08(-0.44, 0.61) | 0.747 | -0.21(-0.69, 0.28) | 0.388 |
| R_OFC | 0.92 | 2,66 | 0.405 | 0.733 | 0.3(-0.25, 0.86) | 0.274 | 0.33(-0.19, 0.86) | 0.203 | 0.03(-0.46, 0.52) | 0.906 |
| R_IFG | 0.03 | 2,67 | 0.972 | 0.998 | -0.01(-0.56, 0.54) | 0.975 | 0.04(-0.48, 0.57) | 0.864 | 0.05(-0.43, 0.54) | 0.826 |
| L_PHG | 1.61 | 2,65 | 0.208 | 0.464 | 0.48(-0.08, 1.04) | 0.089 | 0.17(-0.36, 0.7) | 0.524 | -0.31(-0.8, 0.18) | 0.208 |
| R_PHG | 2.61 | 2,68 | 0.081 | 0.237 | 0.47(-0.08, 1.01) | 0.087 | -0.04(-0.56, 0.48) | 0.883 | -0.5(-0.99, -0.02) | 0.037 |
| L_VC | 0.41 | 2,68 | 0.664 | 0.857 | -0.15(-0.69, 0.4) | 0.587 | 0.07(-0.45, 0.59) | 0.793 | 0.22(-0.27, 0.7) | 0.371 |
| R_VC | 0.32 | 2,66 | 0.728 | 0.892 | 0.13(-0.42, 0.69) | 0.634 | -0.06(-0.58, 0.46) | 0.815 | -0.19(-0.68, 0.3) | 0.43 |
| L_VS | 0 | 2,68 | 0.998 | 0.998 | -0.01(-0.55, 0.53) | 0.967 | -0.02(-0.54, 0.5) | 0.948 | -0.01(-0.49, 0.48) | 0.981 |
| R_VS | 0.48 | 2,66 | 0.623 | 0.849 | -0.24(-0.78, 0.31) | 0.381 | -0.05(-0.57, 0.47) | 0.859 | 0.19(-0.29, 0.68) | 0.427 |
| preSMA | 2.42 | 2,70 | 0.097 | 0.245 | -0.53(-1.07, 0.01) | 0.05 | -0.49(-1.02, 0.03) | 0.058 | 0.03(-0.44, 0.51) | 0.886 |
| rACC | 0.7 | 2,65 | 0.499 | 0.792 | -0.3(-0.85, 0.25) | 0.272 | -0.08(-0.62, 0.45) | 0.749 | 0.22(-0.27, 0.71) | 0.372 |

**Main effect pfdr < 0.05 AND sham vs. active p < 0.05; tMain effect pfdr<010 AND sham vs. active p < 0.05.*
*P values listed as "0" are p < 0.001.*
*CI, 95% Confidence Interval.*
*For pairwise comparisons, negative Cohen's d indicates that first condition listed is less than second condition (e.g., NetStrength is less for sham than active if Cohen's d is negative).*

**Table S10. LTA-CtDCS Dual Regression Connectivity**

| | Main Effect | | | | Sham vs. Active | | Rest vs. Active | | Rest vs. Inactive | |
|---|---|---|---|---|---|---|---|---|---|---|
| roi | F | DF | p | $p_{FDR}$ | Cohen's d (CI) | p | Cohen's d (CI) | p | Cohen's d (CI) | p |
| netStrength | 0.17 | 2,62 | 0.842 | 1 | 0.09(-0.41, 0.58) | 0.728 | -0.08(-0.62, 0.46) | 0.767 | -0.17(-0.74, 0.41) | 0.562 |
| L_HG | 0 | 2,62 | 0.996 | 1 | 0.02(-0.48, 0.52) | 0.934 | 0.01(-0.52, 0.55) | 0.957 | -0.01(-0.57, 0.56) | 0.983 |
| R_HG | 0.24 | 2,62 | 0.788 | 1 | -0.17(-0.67, 0.33) | 0.492 | -0.07(-0.61, 0.47) | 0.794 | 0.1(-0.47, 0.68) | 0.725 |
| L_lHG | 0.6 | 2,62 | 0.552 | 1 | 0.2(-0.31, 0.71) | 0.435 | -0.1(-0.64, 0.44) | 0.702 | -0.3(-0.88, 0.27) | 0.295 |
| R_lHG | 0.82 | 2,62 | 0.445 | 1 | -0.28(-0.78, 0.22) | 0.263 | -0.27(-0.81, 0.27) | 0.321 | 0.01(-0.56, 0.58) | 0.962 |
| L_mHG | 0.38 | 2,62 | 0.685 | 1 | 0.21(-0.29, 0.71) | 0.405 | 0.15(-0.39, 0.69) | 0.584 | -0.06(-0.63, 0.51) | 0.835 |
| R_mHG | 0.44 | 2,61 | 0.649 | 1 | 0.13(-0.37, 0.63) | 0.605 | 0.25(-0.29, 0.78) | 0.363 | 0.12(-0.46, 0.69) | 0.687 |
| L_MGN | 2.76 | 2,62 | 0.071 | 1 | 0.58(0.08, 1.09) | 0.023 | 0.31(-0.23, 0.84) | 0.257 | -0.28(-0.84, 0.29) | 0.337 |
| R_MGN | 0.24 | 2,63 | 0.784 | 1 | 0.05(-0.45, 0.55) | 0.838 | 0.19(-0.35, 0.72) | 0.49 | 0.13(-0.43, 0.7) | 0.637 |
| L_IC | 1.15 | 2,62 | 0.323 | 1 | 0.31(-0.19, 0.8) | 0.225 | 0.35(-0.19, 0.89) | 0.198 | 0.05(-0.52, 0.62) | 0.873 |
| R_IC | 0.05 | 2,63 | 0.947 | 1 | -0.02(-0.52, 0.47) | 0.933 | -0.09(-0.62, 0.45) | 0.745 | -0.07(-0.63, 0.5) | 0.816 |
| L_dCB | 0.23 | 2,57 | 0.794 | 1 | -0.16(-0.67, 0.34) | 0.519 | -0.13(-0.68, 0.43) | 0.65 | 0.04(-0.54, 0.61) | 0.896 |
| R_dCB | 0.09 | 2,62 | 0.919 | 1 | -0.1(-0.6, 0.4) | 0.690 | -0.02(-0.55, 0.52) | 0.945 | 0.08(-0.49, 0.65) | 0.776 |
| L_vCB | 0.07 | 2,62 | 0.929 | 1 | -0.02(-0.51, 0.48) | 0.938 | 0.08(-0.45, 0.62) | 0.754 | 0.1(-0.47, 0.67) | 0.717 |
| R_vCB | 0.53 | 2,63 | 0.592 | 1 | 0.2(-0.29, 0.7) | 0.421 | 0.24(-0.29, 0.78) | 0.371 | 0.04(-0.53, 0.61) | 0.89 |
| L_DN | 0.94 | 2,87 | 0.394 | 1 | 0.2(-0.29, 0.7) | 0.417 | 0.36(-0.18, 0.91) | 0.187 | 0.16(-0.42, 0.73) | 0.585 |
| R_DN | 0.02 | 2,90 | 0.983 | 1 | -0.03(-0.52, 0.47) | 0.907 | 0.02(-0.51, 0.56) | 0.932 | 0.05(-0.51, 0.62) | 0.855 |
| L_dCS | 0 | 2,57 | 1 | 1 | -0.01(-0.51, 0.5) | 0.976 | 0(-0.56, 0.56) | 0.988 | 0(-0.58, 0.58) | 0.99 |
| R_dCS | 0.57 | 2,63 | 0.566 | 1 | 0.27(-0.23, 0.77) | 0.289 | 0.1(-0.44, 0.63) | 0.719 | -0.17(-0.75, 0.41) | 0.554 |
| L_CS | 1.79 | 2,59 | 0.175 | 1 | 0.11(-0.4, 0.62) | 0.664 | -0.41(-0.96, 0.14) | 0.137 | -0.52(-1.1, 0.06) | 0.074 |
| R_CS | 2.07 | 2,59 | 0.136 | 1 | -0.29(-0.81, 0.22) | 0.258 | -0.54(-1.1, 0.01) | 0.051 | -0.25(-0.84, 0.34) | 0.396 |
| L_Th | 3.26 | 2,62 | 0.045 | 1 | 0.35(-0.15, 0.85) | 0.162 | 0.68(0.13, 1.22) | 0.015 | 0.33(-0.25, 0.9) | 0.261 |
| R_Th | 1.52 | 2,62 | 0.226 | 1 | 0.41(-0.09, 0.91) | 0.103 | 0.31(-0.23, 0.85) | 0.254 | -0.1(-0.67, 0.47) | 0.724 |
| L_aIns | 0.16 | 2,63 | 0.856 | 1 | -0.13(-0.63, 0.37) | 0.606 | 0(-0.54, 0.53) | 0.988 | 0.12(-0.44, 0.69) | 0.662 |
| R_aIns | 0.37 | 2,62 | 0.69 | 1 | 0.2(-0.3, 0.69) | 0.433 | 0.17(-0.37, 0.71) | 0.528 | -0.03(-0.6, 0.55) | 0.93 |
| L_DLPFC | 0.81 | 2,60 | 0.451 | 1 | -0.16(-0.66, 0.34) | 0.527 | -0.34(-0.88, 0.2) | 0.212 | -0.18(-0.75, 0.39) | 0.532 |
| R_DLPFC | 0.51 | 2,59 | 0.604 | 1 | -0.08(-0.59, 0.44) | 0.769 | -0.27(-0.82, 0.27) | 0.321 | -0.2(-0.77, 0.38) | 0.496 |
| L_OFC | 0.13 | 2,62 | 0.881 | 1 | 0(-0.5, 0.5) | 0.999 | 0.13(-0.41, 0.66) | 0.643 | 0.13(-0.46, 0.71) | 0.668 |
| R_OFC | 1.18 | 2,61 | 0.315 | 1 | 0.38(-0.12, 0.87) | 0.136 | 0.23(-0.31, 0.76) | 0.405 | -0.15(-0.72, 0.42) | 0.603 |
| R_IFG | 0.15 | 2,61 | 0.86 | 1 | -0.14(-0.64, 0.37) | 0.591 | -0.08(-0.62, 0.45) | 0.762 | 0.05(-0.52, 0.62) | 0.85 |
| L_PHG | 1.23 | 2,63 | 0.299 | 1 | 0.34(-0.16, 0.83) | 0.181 | 0.34(-0.2, 0.87) | 0.211 | 0(-0.57, 0.57) | 0.996 |
| R_PHG | 0.23 | 2,63 | 0.798 | 1 | -0.08(-0.58, 0.41) | 0.737 | 0.11(-0.43, 0.64) | 0.689 | 0.19(-0.37, 0.76) | 0.503 |
| L_VC | 0.47 | 2,62 | 0.63 | 1 | 0.12(-0.39, 0.63) | 0.636 | -0.16(-0.7, 0.39) | 0.571 | -0.28(-0.85, 0.3) | 0.338 |
| R_VC | 0.75 | 2,61 | 0.475 | 1 | 0.01(-0.5, 0.51) | 0.983 | -0.3(-0.84, 0.24) | 0.268 | -0.31(-0.88, 0.27) | 0.289 |
| L_VS | 0.62 | 2,60 | 0.539 | 1 | -0.23(-0.73, 0.28) | 0.371 | -0.27(-0.81, 0.28) | 0.337 | -0.04(-0.61, 0.54) | 0.898 |
| R_VS | 0.43 | 2,61 | 0.651 | 1 | -0.08(-0.58, 0.42) | 0.747 | -0.25(-0.78, 0.29) | 0.356 | -0.17(-0.74, 0.4) | 0.56 |
| preSMA | 0.27 | 2,59 | 0.767 | 1 | -0.07(-0.57, 0.43) | 0.788 | -0.2(-0.75, 0.35) | 0.469 | -0.13(-0.72, 0.45) | 0.65 |
| rACC | 0.21 | 2,61 | 0.815 | 0.9 | 0.08(-0.43, 0.58) | 0.766 | -0.15(-0.69, 0.38) | 0.57 | -0.23(-0.8, 0.34) | 0.426 |

**Main effect pfdr < 0.05 AND sham vs. active p < 0.05; tMain effect pfdr<010 AND sham vs. active p < 0.05.*
*P values listed as "0" are p < 0.001.*
*CI, 95% Confidence Interval.*
*For pairwise comparisons, negative Cohen's d indicates that first condition listed is less than second condition (e.g., NetStrength is less for sham than active if Cohen's d is negative).*

**Table S11. STC-CtDCS Dual Regression Connectivity**

| | Main Effect | | | | Sham vs. Active | | Rest vs. Active | | Rest vs. Inactive | |
|---|---|---|---|---|---|---|---|---|---|---|
| roi | F | DF | p | $p_{FDR}$ | Cohen's d (CI) | p | Cohen's d (CI) | p | Cohen's d (CI) | p |
| netStrength | 5.4 | 2,17 | 0.015 | 0.583 | 1.5(0.33, 2.66) | 0.005 | 0.27(-1.37, 1.91) | 0.71 | -1.23(-3.07, 0.62) | 0.138 |
| L_HG | 1.19 | 2,18 | 0.327 | 0.694 | 0.71(-0.39, 1.81) | 0.156 | -0.11(-1.73, 1.51) | 0.883 | -0.82(-2.68, 1.03) | 0.336 |
| R_HG | 0.36 | 2,17 | 0.705 | 0.931 | 0.41(-0.71, 1.53) | 0.42 | 0.17(-1.54, 1.88) | 0.827 | -0.24(-2.12, 1.64) | 0.78 |
| L_lHG | 0.46 | 2,17 | 0.641 | 0.879 | 0.43(-0.68, 1.54) | 0.388 | -0.18(-1.86, 1.49) | 0.805 | -0.61(-2.53, 1.31) | 0.481 |
| R_lHG | 0.71 | 2,17 | 0.504 | 0.829 | 0.34(-0.7, 1.38) | 0.467 | -0.56(-2.19, 1.07) | 0.445 | -0.9(-2.69, 0.89) | 0.267 |
| L_mHG | 1.97 | 2,18 | 0.169 | 0.694 | 0.89(-0.18, 1.95) | 0.068 | 0.01(-1.58, 1.61) | 0.984 | -0.87(-2.63, 0.88) | 0.278 |
| R_mHG | 1.37 | 2,16 | 0.282 | 0.694 | 0.71(-0.45, 1.87) | 0.169 | 0.67(-1.04, 2.38) | 0.379 | -0.04(-1.98, 1.89) | 0.961 |
| L_MGN | 0.8 | 2,17 | 0.464 | 0.817 | -0.13(-1.16, 0.91) | 0.79 | 0.79(-0.84, 2.41) | 0.291 | 0.91(-0.81, 2.64) | 0.255 |
| R_MGN | 1.3 | 2,22 | 0.292 | 0.694 | 0.35(-0.67, 1.37) | 0.469 | 1.04(-0.53, 2.62) | 0.154 | 0.7(-0.96, 2.35) | 0.37 |
| L_IC | 1.64 | 2,18 | 0.221 | 0.694 | 0.82(-0.24, 1.87) | 0.091 | 0.06(-1.53, 1.65) | 0.934 | -0.76(-2.51, 0.99) | 0.345 |
| R_IC | 1.9 | 2,18 | 0.178 | 0.694 | 0.84(-0.17, 1.86) | 0.083 | 0.65(-0.92, 2.23) | 0.368 | -0.19(-1.89, 1.52) | 0.813 |
| L_dCB | 1.93 | 2,17 | 0.175 | 0.694 | 0.86(-0.21, 1.94) | 0.076 | 0.58(-1.05, 2.21) | 0.43 | -0.29(-2.05, 1.48) | 0.72 |
| R_dCB | 2.1 | 2,16 | 0.155 | 0.694 | 0.42(-0.68, 1.52) | 0.402 | 1.53(-0.3, 3.35) | 0.066 | 1.11(-0.73, 2.94) | 0.185 |
| L_vCB | 0.63 | 2,23 | 0.542 | 0.829 | 0.35(-0.64, 1.33) | 0.456 | 0.64(-0.88, 2.15) | 0.368 | 0.29(-1.33, 1.92) | 0.705 |
| R_vCB | 0.06 | 2,17 | 0.943 | 0.969 | -0.09(-1.1, 0.92) | 0.844 | 0.17(-1.42, 1.75) | 0.818 | 0.26(-1.46, 1.98) | 0.745 |
| L_DN | 1.17 | 2,17 | 0.335 | 0.694 | 0.46(-0.6, 1.52) | 0.338 | -0.65(-2.31, 1.01) | 0.397 | -1.11(-2.85, 0.62) | 0.177 |
| R_DN | 3.87 | 2,17 | 0.041 | 0.694 | 0.14(-0.89, 1.17) | 0.763 | -1.89(-3.52, -0.26) | 0.017 | -2.03(-3.8, -0.25) | 0.019 |
| L_dCS | 0.56 | 2,17 | 0.582 | 0.829 | -0.24(-1.29, 0.81) | 0.626 | -0.77(-2.38, 0.84) | 0.323 | -0.53(-2.19, 1.14) | 0.5 |
| R_dCS | 0.1 | 2,17 | 0.901 | 0.953 | 0.04(-0.99, 1.06) | 0.938 | -0.3(-1.92, 1.32) | 0.677 | -0.34(-2.1, 1.42) | 0.671 |
| L_CS | 0.17 | 2,16 | 0.848 | 0.953 | 0.26(-0.82, 1.33) | 0.593 | 0.21(-1.46, 1.88) | 0.78 | -0.05(-1.83, 1.74) | 0.952 |
| R_CS | 2.61 | 2,17 | 0.103 | 0.694 | 0.92(-0.17, 2.02) | 0.06 | 0.97(-0.71, 2.65) | 0.193 | 0.05(-1.75, 1.85) | 0.949 |
| L_Th | 0.14 | 2,18 | 0.867 | 0.953 | 0.22(-0.8, 1.25) | 0.642 | 0.25(-1.36, 1.87) | 0.737 | 0.03(-1.67, 1.73) | 0.969 |
| R_Th | 3.67 | 2,17 | 0.047 | 0.694 | 0.73(-0.31, 1.77) | 0.13 | 1.72(0.03, 3.4) | 0.028 | 0.99(-0.82, 2.8) | 0.225 |
| L_aIns | 0.02 | 2,17 | 0.983 | 0.983 | -0.07(-1.11, 0.96) | 0.875 | 0.05(-1.58, 1.68) | 0.948 | 0.12(-1.66, 1.9) | 0.88 |
| R_aIns | 1.16 | 2,16 | 0.34 | 0.694 | 0.12(-0.98, 1.22) | 0.808 | -1.18(-2.96, 0.61) | 0.153 | -1.3(-3.36, 0.77) | 0.187 |
| L_DLPFC | 0.65 | 2,16 | 0.535 | 0.829 | 0.53(-0.58, 1.64) | 0.285 | -0.1(-1.76, 1.57) | 0.897 | -0.63(-2.54, 1.28) | 0.467 |
| R_DLPFC | 0.22 | 2,18 | 0.809 | 0.953 | -0.24(-1.27, 0.78) | 0.602 | 0.2(-1.4, 1.8) | 0.785 | 0.44(-1.3, 2.18) | 0.581 |
| L_OFC | 1.05 | 2,18 | 0.371 | 0.694 | -0.04(-1.05, 0.97) | 0.928 | -0.99(-2.59, 0.61) | 0.179 | -0.95(-2.68, 0.78) | 0.238 |
| R_OFC | 1.04 | 2,18 | 0.375 | 0.694 | -0.44(-1.47, 0.6) | 0.351 | -0.85(-2.46, 0.75) | 0.246 | -0.42(-2.16, 1.32) | 0.6 |
| R_IFG | 0.56 | 2,16 | 0.583 | 0.829 | 0.26(-0.8, 1.32) | 0.586 | 0.71(-0.95, 2.37) | 0.344 | 0.45(-1.32, 2.22) | 0.576 |
| L_PHG | 0.2 | 2,23 | 0.823 | 0.953 | -0.19(-1.17, 0.8) | 0.686 | -0.36(-1.87, 1.14) | 0.606 | -0.18(-1.8, 1.45) | 0.818 |
| R_PHG | 1.52 | 2,18 | 0.246 | 0.694 | -0.68(-1.71, 0.35) | 0.154 | 0.43(-1.17, 2.03) | 0.555 | 1.11(-0.65, 2.87) | 0.172 |
| L_VC | 2.88 | 2,23 | 0.077 | 0.694 | 0.86(-0.16, 1.88) | 0.076 | -0.69(-2.19, 0.82) | 0.333 | -1.55(-3.18, 0.08) | 0.054 |
| R_VC | 0.15 | 2,18 | 0.864 | 0.953 | -0.13(-1.13, 0.87) | 0.778 | 0.27(-1.28, 1.82) | 0.707 | 0.4(-1.28, 2.08) | 0.61 |
| L_VS | 0.12 | 2,18 | 0.885 | 0.953 | 0.09(-0.93, 1.11) | 0.845 | -0.29(-1.88, 1.31) | 0.693 | -0.38(-2.11, 1.35) | 0.635 |
| R_VS | 2.46 | 2,23 | 0.108 | 0.694 | 0.61(-0.4, 1.61) | 0.201 | -0.96(-2.46, 0.54) | 0.181 | -1.57(-3.21, 0.08) | 0.051 |
| preSMA | 1.06 | 2,17 | 0.367 | 0.694 | 0.65(-0.38, 1.68) | 0.17 | 0.35(-1.27, 1.97) | 0.631 | -0.3(-2.07, 1.46) | 0.703 |
| rACC | 1.7 | 2,19 | 0.21 | 0.694 | 0.87(-0.22, 1.96) | 0.088 | 0.07(-1.47, 1.61) | 0.922 | -0.8(-2.54, 0.94) | 0.327 |

**Main effect pfdr < 0.05 AND sham vs. active p < 0.05; tMain effect pfdr<010 AND sham vs. active p < 0.05.*
*P values listed as "0" are p < 0.001.*
*CI, 95% Confidence Interval.*
*For pairwise comparisons, negative Cohen's d indicates that first condition listed is less than second condition (e.g., NetStrength is less for active than sham if Cohen's d is positive).*

**Table S12.DLPFC-CtDCS Dual Regression Connectivity**

| | Main Effect | | | | Sham vs. Active | | Rest vs. Active | | Rest vs. Inactive | |
|---|---|---|---|---|---|---|---|---|---|---|
| roi | F | DF | p | $p_{FDR}$ | Cohen's d (CI) | p | Cohen's d (CI) | p | Cohen's d (CI) | p |
| netStrength | 2.85 | 2,49 | 0.067 | 0.366 | 0.28(-0.31, 0.87) | 0.342 | -0.38(-0.95, 0.19) | 0.172 | -0.66(-1.24, -0.08) | 0.022 |
| L_HG | 5.52 | 2,50 | 0.007 | 0.129 | -0.22(-0.81, 0.38) | 0.461 | -0.87(-1.46, -0.28) | 0.003 | -0.66(-1.24, -0.08) | 0.021 |
| R_HG | 1.43 | 2,46 | 0.25 | 0.704 | 0.44(-0.17, 1.05) | 0.143 | 0.02(-0.56, 0.59) | 0.953 | -0.42(-1.02, 0.17) | 0.146 |
| L_lHG | 6.4 | 2,47 | 0.003 | 0.129 | 0.2(-0.41, 0.8) | 0.504 | -0.75(-1.35, -0.16) | 0.01 | -0.95(-1.56, -0.34) | 0.002 |
| R_lHG | 3.29 | 2,48 | 0.046 | 0.366 | 0.75(0.13, 1.36) | 0.014 | 0.42(-0.15, 0.99) | 0.133 | -0.33(-0.91, 0.26) | 0.261 |
| L_mHG | 1.5 | 2,50 | 0.233 | 0.704 | -0.08(-0.65, 0.5) | 0.79 | -0.43(-0.99, 0.12) | 0.116 | -0.36(-0.91, 0.2) | 0.193 |
| R_mHG | 0.63 | 2,51 | 0.534 | 0.932 | 0.21(-0.38, 0.8) | 0.474 | -0.1(-0.66, 0.46) | 0.718 | -0.31(-0.87, 0.26) | 0.27 |
| L_MGN | 1 | 2,52 | 0.376 | 0.834 | 0.39(-0.17, 0.96) | 0.165 | 0.19(-0.35, 0.73) | 0.475 | -0.2(-0.74, 0.34) | 0.461 |
| R_MGN | 1.66 | 2,52 | 0.201 | 0.704 | 0.5(-0.07, 1.07) | 0.082 | 0.34(-0.21, 0.89) | 0.213 | -0.16(-0.7, 0.39) | 0.563 |
| L_IC | 1.18 | 2,53 | 0.316 | 0.751 | 0.35(-0.22, 0.92) | 0.218 | 0.38(-0.17, 0.93) | 0.169 | 0.03(-0.53, 0.59) | 0.922 |
| R_IC | 0.11 | 2,52 | 0.897 | 0.986 | -0.05(-0.63, 0.53) | 0.867 | 0.08(-0.46, 0.62) | 0.778 | 0.12(-0.42, 0.67) | 0.649 |
| L_dCB | 0.24 | 2,54 | 0.784 | 0.955 | -0.16(-0.72, 0.4) | 0.576 | -0.17(-0.7, 0.36) | 0.529 | -0.01(-0.56, 0.54) | 0.972 |
| R_dCB | 0.41 | 2,50 | 0.663 | 0.955 | -0.13(-0.71, 0.44) | 0.647 | -0.25(-0.81, 0.31) | 0.369 | -0.12(-0.68, 0.44) | 0.675 |
| L_vCB | 0.01 | 2,54 | 0.991 | 0.991 | -0.01(-0.57, 0.56) | 0.982 | -0.03(-0.57, 0.51) | 0.903 | -0.03(-0.57, 0.52) | 0.923 |
| R_vCB | 0.04 | 2,52 | 0.965 | 0.991 | 0.07(-0.5, 0.65) | 0.794 | 0.02(-0.52, 0.57) | 0.928 | -0.05(-0.61, 0.5) | 0.855 |
| L_DN | 2.81 | 2,72 | 0.067 | 0.366 | -0.45(-1.02, 0.12) | 0.115 | -0.62(-1.17, -0.07) | 0.025 | -0.17(-0.72, 0.39) | 0.547 |
| R_DN | 0.46 | 2,74 | 0.635 | 0.955 | -0.27(-0.83, 0.29) | 0.345 | -0.13(-0.66, 0.39) | 0.613 | 0.13(-0.42, 0.68) | 0.63 |
| L_dCS | 0.89 | 2,57 | 0.417 | 0.834 | 0.05(-0.51, 0.62) | 0.847 | -0.27(-0.81, 0.26) | 0.306 | -0.33(-0.88, 0.22) | 0.231 |
| R_dCS | 1.38 | 2,54 | 0.259 | 0.704 | 0.23(-0.34, 0.79) | 0.429 | -0.23(-0.76, 0.31) | 0.405 | -0.45(-1, 0.1) | 0.103 |
| L_CS | 2.06 | 2,56 | 0.137 | 0.65 | 0.35(-0.22, 0.92) | 0.215 | -0.18(-0.72, 0.35) | 0.491 | -0.54(-1.08, 0.01) | 0.049 |
| R_CS | 1.26 | 2,54 | 0.293 | 0.741 | 0.43(-0.16, 1.02) | 0.14 | 0.09(-0.46, 0.63) | 0.747 | -0.34(-0.9, 0.22) | 0.216 |
| L_Th | 0.23 | 2,72 | 0.795 | 0.955 | 0.15(-0.42, 0.71) | 0.612 | -0.03(-0.58, 0.51) | 0.906 | -0.18(-0.73, 0.37) | 0.52 |
| R_Th | 0.19 | 2,54 | 0.83 | 0.955 | 0.01(-0.57, 0.58) | 0.982 | -0.14(-0.69, 0.41) | 0.616 | -0.14(-0.69, 0.4) | 0.595 |
| L_aIns | 1.84 | 2,52 | 0.169 | 0.704 | -0.53(-1.1, 0.04) | 0.064 | -0.33(-0.87, 0.22) | 0.235 | 0.21(-0.34, 0.76) | 0.455 |
| R_aIns | 0.29 | 2,54 | 0.749 | 0.955 | -0.06(-0.62, 0.51) | 0.844 | -0.2(-0.74, 0.34) | 0.466 | -0.14(-0.69, 0.41) | 0.609 |
| L_DLPFC | 0.22 | 2,71 | 0.802 | 0.955 | -0.14(-0.72, 0.43) | 0.624 | 0.04(-0.5, 0.57) | 0.888 | 0.18(-0.38, 0.74) | 0.524 |
| R_DLPFC | 1.54 | 2,55 | 0.224 | 0.704 | 0.38(-0.18, 0.95) | 0.179 | 0.44(-0.1, 0.99) | 0.106 | 0.06(-0.48, 0.6) | 0.821 |
| L_OFC | 0.38 | 2,53 | 0.688 | 0.955 | -0.13(-0.69, 0.43) | 0.64 | -0.23(-0.78, 0.31) | 0.391 | -0.1(-0.64, 0.44) | 0.705 |
| R_OFC | 0.31 | 2,53 | 0.736 | 0.955 | 0.19(-0.38, 0.75) | 0.508 | 0.19(-0.35, 0.73) | 0.488 | 0(-0.54, 0.54) | 0.996 |
| R_IFG | 4.54 | 2,52 | 0.015 | 0.192 | -0.47(-1.06, 0.12) | 0.111 | -0.84(-1.41, -0.27) | 0.004 | -0.37(-0.92, 0.18) | 0.185 |
| L_PHG | 0.62 | 2,54 | 0.54 | 0.932 | -0.05(-0.62, 0.51) | 0.848 | -0.28(-0.83, 0.27) | 0.307 | -0.23(-0.77, 0.31) | 0.402 |
| R_PHG | 0.94 | 2,47 | 0.396 | 0.834 | 0.27(-0.3, 0.84) | 0.347 | 0.37(-0.19, 0.92) | 0.187 | 0.1(-0.46, 0.65) | 0.727 |
| L_VC | 0.03 | 2,48 | 0.967 | 0.991 | -0.06(-0.63, 0.5) | 0.828 | -0.06(-0.6, 0.47) | 0.816 | 0(-0.54, 0.54) | 0.997 |
| R_VC | 0.52 | 2,50 | 0.6 | 0.955 | 0.18(-0.39, 0.76) | 0.523 | -0.09(-0.65, 0.47) | 0.739 | -0.27(-0.83, 0.28) | 0.321 |
| L_VS | 0.1 | 2,73 | 0.909 | 0.986 | -0.01(-0.58, 0.56) | 0.971 | -0.1(-0.63, 0.43) | 0.694 | -0.09(-0.64, 0.46) | 0.733 |
| R_VS | 0.19 | 2,72 | 0.825 | 0.955 | 0.09(-0.48, 0.65) | 0.763 | -0.08(-0.62, 0.46) | 0.758 | -0.17(-0.72, 0.38) | 0.539 |
| preSMA | 3.21 | 2,49 | 0.049 | 0.366 | 0.68(0.11, 1.26) | 0.018 | 0.49(-0.06, 1.04) | 0.074 | -0.2(-0.75, 0.36) | 0.478 |
| rACC | 0.7 | 2,53 | 0.502 | 0.932 | -0.18(-0.74, 0.39) | 0.532 | 0.14(-0.38, 0.67) | 0.585 | 0.32(-0.23, 0.87) | 0.243 |

**Main effect pfdr < 0.05 AND sham vs. active p < 0.05; tMain effect pfdr<010 AND sham vs. active p < 0.05.*
*P values listed as "0" are p < 0.001.*
*CI, 95% Confidence Interval.*
*For pairwise comparisons, negative Cohen's d indicates that first condition listed is less than second condition (e.g., NetStrength is less for active than sham if Cohen's d is positive).*

**Table S13.Langguth-4x1tDCS Targeting Auditory Cortex, HG/HS Seed Connectivity**

| | Interaction, Condition-by-Time | | | | Active, Session 1 vs. Session 2 | | Sham, Session 1 vs. Session 2 | |
|---|---|---|---|---|---|---|---|---|
| roi | t | DF | p | $p_{FDR}$ | Cohen’s d (CI) | p | Cohen’s d (CI) | p |
| netStrength | -0.03 | 15 | 0.973 | 0.973 | -0.89(-1.9, 0.12) | 0.078 | -0.87(-1.87, 0.14) | 0.083 |
| L_HG | 0.66 | 17 | 0.515 | 0.926 | 0.12(-0.87, 1.1) | 0.810 | -0.32(-1.25, 0.62) | 0.489 |
| R_HG | -0.49 | 17 | 0.634 | 0.926 | -0.75(-1.75, 0.25) | 0.131 | -0.43(-1.37, 0.5) | 0.346 |
| L_lHG | -0.52 | 17 | 0.609 | 0.926 | -0.4(-1.37, 0.57) | 0.406 | -0.06(-0.98, 0.85) | 0.89 |
| R_lHG | -0.38 | 15 | 0.707 | 0.959 | -0.44(-1.43, 0.55) | 0.364 | -0.19(-1.16, 0.79) | 0.696 |
| L_mHG | 0.16 | 15 | 0.875 | 0.959 | -0.59(-1.6, 0.42) | 0.227 | -0.7(-1.72, 0.32) | 0.158 |
| R_mHG | -2.7 | 16 | 0.016 | 0.591 | -1.58(-2.67, -0.49) | 0.004 | 0.22(-0.78, 1.22) | 0.65 |
| L_MGN | 0.84 | 16 | 0.414 | 0.926 | 0.28(-0.69, 1.25) | 0.564 | -0.28(-1.23, 0.67) | 0.559 |
| R_MGN | -0.79 | 15 | 0.443 | 0.926 | 0.08(-0.89, 1.05) | 0.861 | 0.61(-0.4, 1.63) | 0.224 |
| L_IC | -0.64 | 16 | 0.53 | 0.926 | -0.33(-1.31, 0.66) | 0.498 | 0.1(-0.88, 1.08) | 0.836 |
| R_IC | 1.28 | 14 | 0.222 | 0.903 | 0.65(-0.34, 1.64) | 0.190 | -0.22(-1.23, 0.79) | 0.661 |
| L_dCB | 1.37 | 15 | 0.19 | 0.903 | -0.36(-1.35, 0.62) | 0.454 | -1.25(-2.22, -0.29) | 0.012 |
| R_dCB | 1.75 | 17 | 0.098 | 0.903 | 0.32(-0.66, 1.3) | 0.511 | -0.82(-1.75, 0.1) | 0.085 |
| L_vCB | -0.52 | 16 | 0.608 | 0.926 | -1.29(-2.34, -0.25) | 0.014 | -0.95(-1.92, 0.02) | 0.049 |
| R_vCB | 0.1 | 16 | 0.918 | 0.959 | -0.75(-1.74, 0.24) | 0.133 | -0.82(-1.85, 0.21) | 0.113 |
| L_DN | -1.88 | 28 | 0.071 | 0.898 | -0.63(-1.65, 0.38) | 0.213 | 0.64(-0.35, 1.62) | 0.196 |
| R_DN | 0.24 | 15 | 0.815 | 0.959 | -0.5(-1.48, 0.48) | 0.304 | -0.66(-1.64, 0.32) | 0.177 |
| L_dCS | 0.57 | 16 | 0.578 | 0.926 | 0.05(-0.93, 1.03) | 0.921 | -0.32(-1.25, 0.61) | 0.482 |
| R_dCS | 1.23 | 16 | 0.238 | 0.903 | 0.22(-0.77, 1.2) | 0.652 | -0.58(-1.52, 0.36) | 0.213 |
| L_CS | 1.16 | 16 | 0.262 | 0.905 | 0.33(-0.66, 1.31) | 0.500 | -0.43(-1.37, 0.5) | 0.35 |
| R_CS | 0.11 | 17 | 0.916 | 0.959 | -0.05(-1.02, 0.92) | 0.918 | -0.12(-1.04, 0.81) | 0.794 |
| L_Th | -0.1 | 16 | 0.92 | 0.959 | -0.16(-1.14, 0.82) | 0.737 | -0.09(-1.02, 0.83) | 0.835 |
| R_Th | 0.68 | 16 | 0.509 | 0.926 | -0.11(-1.12, 0.89) | 0.818 | -0.56(-1.49, 0.36) | 0.227 |
| L_aIns | -0.45 | 13 | 0.66 | 0.928 | -0.55(-1.54, 0.45) | 0.264 | -0.25(-1.24, 0.74) | 0.608 |
| R_aIns | -1.36 | 14 | 0.195 | 0.903 | -1.23(-2.27, -0.19) | 0.02 | -0.32(-1.31, 0.67) | 0.506 |
| L_DLPFC | 0.65 | 17 | 0.523 | 0.926 | -0.2(-1.18, 0.78) | 0.679 | -0.62(-1.57, 0.32) | 0.183 |
| R_DLPFC | -0.08 | 15 | 0.933 | 0.959 | -0.4(-1.38, 0.59) | 0.410 | -0.34(-1.31, 0.63) | 0.475 |
| L_OFC | -0.95 | 15 | 0.359 | 0.926 | -0.26(-1.3, 0.78) | 0.613 | 0.39(-0.6, 1.38) | 0.423 |
| R_OFC | -2.4 | 14 | 0.031 | 0.591 | -0.38(-1.42, 0.66) | 0.453 | 1.25(0.2, 2.3) | 0.018 |
| R_IFG | 0.29 | 14 | 0.774 | 0.959 | 0.21(-0.84, 1.25) | 0.684 | 0.01(-0.99, 1) | 0.991 |
| L_PHG | 1.31 | 13 | 0.212 | 0.903 | 0.53(-0.49, 1.56) | 0.293 | -0.36(-1.34, 0.62) | 0.46 |
| R_PHG | -1.28 | 15 | 0.218 | 0.903 | 0.18(-0.83, 1.19) | 0.716 | 1.04(0.05, 2.04) | 0.039 |
| L_VC | -0.7 | 17 | 0.496 | 0.926 | -0.44(-1.43, 0.54) | 0.360 | 0.02(-0.95, 0.99) | 0.97 |
| R_VC | 1.03 | 29 | 0.312 | 0.926 | -0.34(-1.31, 0.63) | 0.482 | -1.03(-2.03, -0.02) | 0.045 |
| L_VS | -0.34 | 16 | 0.739 | 0.959 | -0.45(-1.45, 0.56) | 0.360 | -0.22(-1.23, 0.79) | 0.652 |
| R_VS | -0.63 | 14 | 0.542 | 0.926 | -1.08(-2.16, 0) | 0.046 | -0.65(-1.66, 0.35) | 0.188 |
| preSMA | 0.24 | 17 | 0.81 | 0.959 | -0.42(-1.41, 0.57) | 0.389 | -0.58(-1.57, 0.41) | 0.233 |
| rACC | -0.52 | 14 | 0.614 | 0.926 | -0.35(-1.34, 0.64) | 0.474 | 0(-0.99, 0.98) | 0.995 |

**Interaction pfdr < 0.05 AND active session 1 vs. 2 p < 0.05*
*CI, 95% Confidence Interval*
*For pairwise comparisons, negative Cohen's d indicates that first condition listed is less than second condition(e.g., RmHG increases after active tDCS and Cohen's d is negative).*

**Table S14. sgACCnet-4x1tDCS Targeting Left DLPFC, HG/HS Seed Connectivity**

| | Interaction, Condition-by-Time | | | | Active, Session 1 vs. Session 2 | | Sham, Session 1 vs. Session 2 | |
|---|---|---|---|---|---|---|---|---|
| roi | t | DF | p | $p_{FDR}$ | Cohen's d (CI) | p | Cohen's d (CI) | p |
| netStrength | 1.08 | 19 | 0.294 | 0.922 | 0.25(-0.6, 1.09) | 0.553 | -0.42(-1.42, 0.58) | 0.386 |
| L_HG | 1.93 | 21 | 0.067 | 0.922 | 0.34(-0.48, 1.15) | 0.404 | -0.84(-1.83, 0.15) | 0.089 |
| R_HG | 1.51 | 20 | 0.147 | 0.922 | 0.48(-0.34, 1.29) | 0.24 | -0.44(-1.42, 0.54) | 0.358 |
| L_lHG | 0.05 | 20 | 0.961 | 0.985 | 0.54(-0.3, 1.39) | 0.196 | 0.51(-0.42, 1.45) | 0.266 |
| R_lHG | 0.62 | 20 | 0.54 | 0.922 | 0.44(-0.37, 1.26) | 0.274 | 0.06(-0.92, 1.04) | 0.898 |
| L_mHG | 0.88 | 21 | 0.389 | 0.922 | 0.14(-0.7, 0.98) | 0.738 | -0.39(-1.34, 0.55) | 0.395 |
| R_mHG | 0.56 | 21 | 0.579 | 0.922 | 0.09(-0.72, 0.9) | 0.823 | -0.25(-1.18, 0.69) | 0.593 |
| L_MGN | 1.23 | 21 | 0.231 | 0.922 | 0.31(-0.49, 1.11) | 0.438 | -0.42(-1.32, 0.49) | 0.355 |
| R_MGN | -0.2 | 21 | 0.845 | 0.926 | -0.25(-1.05, 0.55) | 0.536 | -0.13(-1.06, 0.81) | 0.782 |
| L_IC | -1.09 | 20 | 0.287 | 0.922 | -0.08(-0.88, 0.72) | 0.837 | 0.58(-0.38, 1.54) | 0.226 |
| R_IC | 0.48 | 22 | 0.638 | 0.922 | 0.41(-0.39, 1.21) | 0.305 | 0.13(-0.81, 1.06) | 0.788 |
| L_dCB | -1.01 | 21 | 0.322 | 0.922 | 0.13(-0.67, 0.93) | 0.75 | 0.74(-0.22, 1.7) | 0.124 |
| R_dCB | 0.02 | 20 | 0.985 | 0.985 | 0(-0.83, 0.84) | 0.991 | -0.01(-0.98, 0.97) | 0.988 |
| L_vCB | 1.09 | 20 | 0.287 | 0.922 | 0.21(-0.6, 1.01) | 0.606 | -0.46(-1.42, 0.5) | 0.338 |
| R_vCB | 0.47 | 21 | 0.642 | 0.922 | 0.09(-0.75, 0.92) | 0.835 | -0.2(-1.12, 0.72) | 0.664 |
| L_DN | -0.37 | 19 | 0.714 | 0.922 | 0.18(-0.66, 1.01) | 0.67 | 0.4(-0.56, 1.37) | 0.398 |
| R_DN | -1.94 | 22 | 0.065 | 0.922 | -0.73(-1.55, 0.09) | 0.078 | 0.45(-0.51, 1.41) | 0.348 |
| L_dCS | -0.95 | 20 | 0.353 | 0.922 | -0.93(-1.82, -0.03) | 0.041 | -0.34(-1.26, 0.58) | 0.449 |
| R_dCS | -1.6 | 16 | 0.129 | 0.922 | -1.08(-1.99, -0.16) | 0.02 | -0.09(-1.03, 0.84) | 0.843 |
| L_CS | -0.96 | 20 | 0.35 | 0.922 | -0.69(-1.55, 0.16) | 0.107 | -0.11(-1.06, 0.85) | 0.821 |
| R_CS | -0.7 | 19 | 0.492 | 0.922 | -0.19(-1.03, 0.65) | 0.654 | 0.24(-0.7, 1.19) | 0.605 |
| L_Th | 0.63 | 21 | 0.537 | 0.922 | 0.41(-0.43, 1.24) | 0.329 | 0.03(-0.87, 0.94) | 0.943 |
| R_Th | -0.68 | 19 | 0.507 | 0.922 | 0.03(-0.83, 0.89) | 0.939 | 0.44(-0.47, 1.36) | 0.332 |
| L_aIns | 1.37 | 21 | 0.186 | 0.922 | 0.63(-0.2, 1.45) | 0.127 | -0.19(-1.13, 0.76) | 0.682 |
| R_aIns | 0.85 | 19 | 0.406 | 0.922 | 0.42(-0.44, 1.28) | 0.324 | -0.09(-1, 0.82) | 0.838 |
| L_DLPFC | -0.32 | 21 | 0.752 | 0.922 | -0.21(-1.07, 0.65) | 0.623 | -0.01(-0.94, 0.91) | 0.975 |
| R_DLPFC | -0.22 | 22 | 0.826 | 0.926 | 0.37(-0.43, 1.17) | 0.358 | 0.5(-0.41, 1.41) | 0.272 |
| L_OFC | -1.39 | 22 | 0.18 | 0.922 | -0.82(-1.64, 0.01) | 0.05 | 0(-0.9, 0.91) | 0.995 |
| R_OFC | -0.38 | 20 | 0.708 | 0.922 | -0.15(-0.95, 0.65) | 0.706 | 0.07(-0.84, 0.99) | 0.869 |
| R_IFG | -0.19 | 22 | 0.853 | 0.926 | -0.35(-1.17, 0.47) | 0.397 | -0.24(-1.14, 0.66) | 0.597 |
| L_PHG | -0.76 | 21 | 0.454 | 0.922 | -0.04(-0.83, 0.76) | 0.926 | 0.41(-0.5, 1.32) | 0.361 |
| R_PHG | 0.34 | 21 | 0.735 | 0.922 | 0.57(-0.25, 1.39) | 0.164 | 0.36(-0.62, 1.34) | 0.455 |
| L_VC | 0.48 | 22 | 0.638 | 0.922 | 0.25(-0.55, 1.05) | 0.534 | -0.03(-0.94, 0.87) | 0.941 |
| R_VC | 1.83 | 22 | 0.082 | 0.922 | 0.63(-0.19, 1.44) | 0.124 | -0.45(-1.37, 0.47) | 0.322 |
| L_VS | 0.39 | 20 | 0.699 | 0.922 | 0.64(-0.2, 1.49) | 0.129 | 0.41(-0.51, 1.33) | 0.373 |
| R_VS | 0.25 | 21 | 0.807 | 0.926 | 0.58(-0.26, 1.43) | 0.165 | 0.44(-0.49, 1.36) | 0.34 |
| preSMA | 0.15 | 20 | 0.882 | 0.931 | -0.17(-1, 0.65) | 0.673 | -0.26(-1.17, 0.64) | 0.56 |
| rACC | 0.76 | 21 | 0.457 | 0.922 | 0.18(-0.62, 0.97) | 0.659 | -0.27(-1.18, 0.63) | 0.546 |

**Interaction pfdr < 0.05 AND active session 1 vs. 2 p < 0.05*
*CI, 95% Confidence Interval*
*For pairwise comparisons, negative Cohen's d indicates that first condition listed is less than second condition(e.g., RmHG increases after active tDCS if Cohen's d is negative).*

**Table S15.Langguth-4x1tDCS Targeting Auditory Cortex, Dual Regression Connectivity**

| | Interaction, Condition-by-Time | | | | Active, Session 1 vs. Session 2 | | Sham, Session 1 vs. Session 2 | |
|---|---|---|---|---|---|---|---|---|
| roi | t | DF | p | $p_{FDR}$ | Cohen's d (CI) | p | Cohen's d (CI) | p |
| netStrength | -1.58 | 14 | 0.136 | 0.527 | -0.74(-1.73, 0.25) | 0.138 | 0.31(-0.65, 1.27) | 0.515 |
| L_HG | -1.07 | 17 | 0.3 | 0.693 | -0.61(-1.61, 0.39) | 0.214 | 0.09(-0.85, 1.02) | 0.851 |
| R_HG | -3.11 | 17 | 0.007 | 0.193 | -0.92(-1.93, 0.1) | 0.069 | 1.1(0.12, 2.08) | 0.025 |
| L_lHG | -2.07 | 17 | 0.054 | 0.497 | -1.06(-2.08, -0.05) | 0.038 | 0.28(-0.65, 1.21) | 0.536 |
| R_lHG | -1.72 | 17 | 0.103 | 0.527 | -0.9(-1.91, 0.11) | 0.073 | 0.22(-0.71, 1.15) | 0.636 |
| L_mHG | -1.61 | 17 | 0.126 | 0.527 | -1.01(-2.05, 0.02) | 0.047 | 0.03(-0.91, 0.98) | 0.942 |
| R_mHG | -2.9 | 17 | 0.01 | 0.193 | -1.17(-2.19, -0.15) | 0.024 | 0.72(-0.22, 1.66) | 0.128 |
| L_MGN | -1.56 | 16 | 0.139 | 0.527 | -0.24(-1.22, 0.73) | 0.613 | 0.8(-0.2, 1.8) | 0.111 |
| R_MGN | -1.79 | 15 | 0.094 | 0.527 | -0.23(-1.21, 0.76) | 0.634 | 0.93(-0.03, 1.9) | 0.053 |
| L_IC | -1.97 | 16 | 0.065 | 0.497 | -0.49(-1.47, 0.48) | 0.311 | 0.79(-0.14, 1.72) | 0.096 |
| R_IC | 0.78 | 17 | 0.447 | 0.699 | 0.12(-0.84, 1.09) | 0.801 | -0.38(-1.31, 0.54) | 0.402 |
| L_dCB | -0.52 | 17 | 0.613 | 0.773 | -0.26(-1.23, 0.7) | 0.583 | 0.07(-0.84, 0.99) | 0.875 |
| R_dCB | -0.2 | 17 | 0.846 | 0.893 | -0.16(-1.14, 0.83) | 0.739 | -0.03(-0.97, 0.9) | 0.944 |
| L_vCB | -0.73 | 17 | 0.475 | 0.699 | -0.83(-1.83, 0.16) | 0.095 | -0.36(-1.28, 0.57) | 0.434 |
| R_vCB | 0.16 | 17 | 0.878 | 0.902 | -0.22(-1.2, 0.75) | 0.64 | -0.33(-1.25, 0.6) | 0.477 |
| L_DN | -0.78 | 17 | 0.446 | 0.699 | -0.19(-1.16, 0.77) | 0.689 | 0.31(-0.6, 1.23) | 0.492 |
| R_DN | 0.62 | 16 | 0.542 | 0.711 | -0.02(-1, 0.96) | 0.968 | -0.42(-1.36, 0.51) | 0.357 |
| L_dCS | 0.88 | 16 | 0.391 | 0.693 | -0.19(-1.17, 0.79) | 0.692 | -0.76(-1.71, 0.19) | 0.107 |
| R_dCS | -0.86 | 15 | 0.401 | 0.693 | 0.01(-0.97, 0.99) | 0.981 | 0.58(-0.41, 1.57) | 0.23 |
| L_CS | 1.08 | 17 | 0.295 | 0.693 | -0.2(-1.18, 0.79) | 0.684 | -0.9(-1.86, 0.07) | 0.062 |
| R_CS | 0.73 | 14 | 0.478 | 0.699 | 0.08(-0.92, 1.07) | 0.874 | -0.41(-1.41, 0.6) | 0.4 |
| L_Th | -0.2 | 30 | 0.842 | 0.893 | 0.15(-0.82, 1.11) | 0.757 | 0.28(-0.67, 1.23) | 0.552 |
| R_Th | -1.44 | 16 | 0.17 | 0.587 | -0.02(-0.99, 0.95) | 0.967 | 0.93(-0.04, 1.9) | 0.063 |
| L_aIns | -1.02 | 17 | 0.321 | 0.693 | -0.74(-1.73, 0.25) | 0.134 | -0.08(-1, 0.84) | 0.863 |
| R_aIns | -0.64 | 12 | 0.533 | 0.711 | -0.68(-1.65, 0.29) | 0.169 | -0.26(-1.21, 0.7) | 0.588 |
| L_DLPFC | 2.12 | 17 | 0.049 | 0.497 | 0.51(-0.48, 1.49) | 0.299 | -0.87(-1.83, 0.08) | 0.068 |
| R_DLPFC | -0.86 | 16 | 0.401 | 0.693 | 0.11(-0.86, 1.08) | 0.823 | 0.68(-0.29, 1.65) | 0.167 |
| L_OFC | 0.04 | 31 | 0.968 | 0.968 | -0.25(-1.21, 0.72) | 0.607 | -0.27(-1.19, 0.64) | 0.55 |
| R_OFC | 0.65 | 31 | 0.523 | 0.711 | -0.11(-1.07, 0.85) | 0.822 | -0.53(-1.44, 0.39) | 0.255 |
| R_IFG | 1.35 | 15 | 0.197 | 0.624 | 0.12(-0.85, 1.1) | 0.797 | -0.75(-1.7, 0.19) | 0.111 |
| L_PHG | 0.34 | 17 | 0.736 | 0.823 | 0.37(-0.62, 1.36) | 0.447 | 0.14(-0.79, 1.08) | 0.75 |
| R_PHG | -0.94 | 31 | 0.354 | 0.693 | -0.17(-1.13, 0.79) | 0.724 | 0.44(-0.47, 1.36) | 0.336 |
| L_VC | -0.43 | 14 | 0.674 | 0.8 | -1.59(-2.67, -0.51) | 0.004 | -1.3(-2.35, -0.26) | 0.013 |
| R_VC | -0.49 | 16 | 0.631 | 0.773 | -0.86(-1.85, 0.14) | 0.088 | -0.54(-1.47, 0.39) | 0.247 |
| L_VS | -0.96 | 16 | 0.351 | 0.693 | -0.39(-1.37, 0.59) | 0.424 | 0.26(-0.76, 1.29) | 0.599 |
| R_VS | -0.39 | 30 | 0.703 | 0.809 | -0.32(-1.29, 0.65) | 0.508 | -0.06(-1.01, 0.88) | 0.891 |
| preSMA | 1.09 | 31 | 0.283 | 0.693 | 0.05(-0.91, 1.02) | 0.91 | -0.66(-1.59, 0.27) | 0.161 |
| rACC | 0.94 | 17 | 0.358 | 0.693 | 0.03(-0.93, 1) | 0.948 | -0.58(-1.51, 0.35) | 0.21 |

**Interaction pfdr < 0.05 AND active session 1 vs. 2 p < 0.05*
*CI, 95% Confidence Interval*
*For pairwise comparisons, negative Cohen's d indicates that first condition listed is less than second condition(e.g., RmHG increases after active tDCS and Cohen's d is negative).*

**Table S16. sgACC-4x1tDCS Targeting Left DLPFC, Dual Regression Connectivity**

| | Interaction, Condition-by-Time | | | | Active, Session 1 vs. Session 2 | | Sham, Session 1 vs. Session 2 | |
|---|---|---|---|---|---|---|---|---|
| roi | t | DF | p | $p_{FDR}$ | Cohen's d (CI) | p | Cohen's d (CI) | p |
| netStrength | -1.21 | 21 | 0.239 | 0.723 | -0.72(-1.58, 0.14) | 0.094 | 0.01(-0.93, 0.95) | 0.979 |
| L_HG | -0.42 | 21 | 0.677 | 0.857 | -0.02(-0.85, 0.81) | 0.963 | 0.23(-0.65, 1.11) | 0.598 |
| R_HG | -0.33 | 19 | 0.745 | 0.913 | -0.06(-0.9, 0.77) | 0.878 | 0.13(-0.78, 1.05) | 0.766 |
| L_lHG | -0.86 | 20 | 0.401 | 0.761 | 0.29(-0.59, 1.16) | 0.506 | 0.8(-0.11, 1.71) | 0.077 |
| R_lHG | -0.71 | 17 | 0.486 | 0.815 | -0.43(-1.28, 0.43) | 0.318 | 0(-0.88, 0.87) | 0.998 |
| L_mHG | 1.03 | 18 | 0.319 | 0.723 | -0.04(-0.91, 0.84) | 0.931 | -0.67(-1.62, 0.28) | 0.155 |
| R_mHG | 0.95 | 21 | 0.354 | 0.723 | 0.12(-0.72, 0.96) | 0.779 | -0.44(-1.34, 0.45) | 0.318 |
| L_MGN | -0.09 | 22 | 0.927 | 0.979 | -0.03(-0.83, 0.77) | 0.934 | 0.02(-0.86, 0.9) | 0.963 |
| R_MGN | 0.73 | 22 | 0.474 | 0.815 | 0.21(-0.61, 1.03) | 0.608 | -0.22(-1.09, 0.65) | 0.616 |
| L_IC | 0.45 | 17 | 0.661 | 0.857 | 0.75(-0.11, 1.62) | 0.085 | 0.49(-0.39, 1.37) | 0.269 |
| R_IC | -1.33 | 21 | 0.198 | 0.723 | -0.17(-0.99, 0.66) | 0.687 | 0.62(-0.28, 1.51) | 0.166 |
| L_dCB | -1.33 | 39 | 0.191 | 0.723 | -0.24(-1.05, 0.58) | 0.56 | 0.54(-0.33, 1.42) | 0.219 |
| R_dCB | -0.03 | 21 | 0.979 | 0.979 | 0.09(-0.75, 0.92) | 0.835 | 0.1(-0.82, 1.02) | 0.822 |
| L_vCB | 0.04 | 16 | 0.972 | 0.979 | 0.16(-0.71, 1.02) | 0.719 | 0.13(-0.74, 1) | 0.758 |
| R_vCB | -0.96 | 38 | 0.343 | 0.723 | -0.04(-0.86, 0.78) | 0.923 | 0.53(-0.37, 1.43) | 0.241 |
| L_DN | 0.6 | 20 | 0.557 | 0.815 | 0.52(-0.31, 1.34) | 0.215 | 0.16(-0.75, 1.07) | 0.731 |
| R_DN | -0.96 | 20 | 0.35 | 0.723 | 0.32(-0.52, 1.15) | 0.447 | 0.89(-0.06, 1.84) | 0.06 |
| L_dCS | -1.38 | 19 | 0.183 | 0.723 | -0.33(-1.17, 0.51) | 0.423 | 0.48(-0.41, 1.38) | 0.274 |
| R_dCS | -2.12 | 22 | 0.046 | 0.723 | -0.84(-1.67, -0.02) | 0.043 | 0.38(-0.5, 1.27) | 0.381 |
| L_CS | -0.51 | 18 | 0.616 | 0.836 | -0.39(-1.27, 0.49) | 0.368 | -0.09(-0.98, 0.8) | 0.843 |
| R_CS | -0.63 | 21 | 0.537 | 0.815 | -0.27(-1.1, 0.57) | 0.519 | 0.1(-0.78, 0.99) | 0.815 |
| L_Th | -1.68 | 22 | 0.107 | 0.723 | -0.87(-1.72, -0.02) | 0.045 | 0.12(-0.76, 0.99) | 0.785 |
| R_Th | -2.33 | 20 | 0.03 | 0.723 | -0.31(-1.15, 0.53) | 0.454 | 1.06(0.14, 1.98) | 0.022 |
| L_aIns | -0.54 | 22 | 0.594 | 0.836 | -0.04(-0.87, 0.79) | 0.93 | 0.28(-0.6, 1.17) | 0.518 |
| R_aIns | -0.94 | 20 | 0.357 | 0.723 | -0.44(-1.28, 0.4) | 0.295 | 0.12(-0.77, 1) | 0.788 |
| L_DLPFC | 0.66 | 21 | 0.513 | 0.815 | 0.07(-0.75, 0.89) | 0.864 | -0.32(-1.2, 0.56) | 0.463 |
| R_DLPFC | -1.13 | 21 | 0.27 | 0.723 | 0.13(-0.72, 0.98) | 0.757 | 0.82(-0.1, 1.75) | 0.08 |
| L_OFC | -1.14 | 22 | 0.267 | 0.723 | -0.6(-1.41, 0.21) | 0.143 | 0.06(-0.81, 0.94) | 0.881 |
| R_OFC | -1.49 | 38 | 0.143 | 0.723 | -0.68(-1.52, 0.15) | 0.112 | 0.2(-0.66, 1.07) | 0.638 |
| R_IFG | 0.06 | 19 | 0.949 | 0.979 | 0.2(-0.62, 1.02) | 0.625 | 0.16(-0.74, 1.06) | 0.718 |
| L_PHG | -0.06 | 17 | 0.955 | 0.979 | -0.02(-0.86, 0.82) | 0.955 | 0.01(-0.89, 0.91) | 0.98 |
| R_PHG | -1.19 | 21 | 0.247 | 0.723 | -0.64(-1.5, 0.22) | 0.138 | 0.07(-0.8, 0.94) | 0.871 |
| L_VC | -2 | 21 | 0.059 | 0.723 | -0.21(-1.02, 0.59) | 0.591 | 1(0.01, 1.99) | 0.044 |
| R_VC | -0.09 | 21 | 0.932 | 0.979 | 0.13(-0.69, 0.95) | 0.752 | 0.18(-0.7, 1.06) | 0.679 |
| L_VS | -0.94 | 21 | 0.358 | 0.723 | -0.55(-1.39, 0.29) | 0.192 | 0(-0.87, 0.88) | 0.994 |
| R_VS | -0.64 | 20 | 0.532 | 0.815 | -0.36(-1.2, 0.49) | 0.393 | 0.02(-0.87, 0.9) | 0.968 |
| preSMA | -0.93 | 22 | 0.362 | 0.723 | -0.17(-0.97, 0.63) | 0.666 | 0.37(-0.51, 1.25) | 0.4 |
| rACC | 0.15 | 38 | 0.88 | 0.979 | 0.31(-0.51, 1.14) | 0.447 | 0.22(-0.67, 1.12) | 0.616 |

**Interaction pfdr < 0.05 AND active session 1 vs. 2 p < 0.05*
*CI, 95% Confidence Interval*
*For pairwise comparisons, negative Cohen's d indicates that first condition listed is less than second condition (e.g., netStrength increases after active tDCS and Cohen's d is negative).*
